# Differential behaviour of a risk score for emergency hospital admission by demographics in Scotland — a retrospective study

**DOI:** 10.1101/2024.02.13.24302753

**Authors:** Ioanna Thoma, Simon Rogers, Jill Ireland, Rachel Porteous, Katie Borland, Catalina A. Vallejos, Louis J. M. Aslett, James Liley

## Abstract

The Scottish Patients at Risk of Re-Admission and Admission (SPARRA) score predicts individual risk of emergency hospital admission for approximately 80% of the Scottish population. It was developed using routinely collected electronic health records, and is used by primary care practitioners to inform anticipatory care, particularly for individuals with high healthcare needs. We comprehensively assess the SPARRA score across population subgroups defined by age, sex, ethnicity, socioeconomic deprivation, and geographic location. For these subgroups, we consider differences in overall performance, score distribution, and false positive and negative rates, using causal methods to identify effects mediated through age, sex, and deprivation. We show that the score is well-calibrated across subgroups, but that rates of false positives and negatives vary widely, mediated by a range of causes. Our work assists practitioners in the application and interpretation of the SPARRA score in population subgroups.

**Research in context:** *Evidence before this study:* There is considerable literature on the general topic of differential performance of risk scores across population subgroups and its implications. A shared theme is the importance of identifying and quantifying such differential performance. We performed a MedLine and Google Scholar search with the single term ‘SPARRA’, and consulted colleagues at Public Health Scotland about any previous internal analyses. Several articles assessed the accuracy of SPARRA and discussed its role in the Scottish healthcare system since its introduction in 2006, but none looked in detail at differential performance between specific demographic groups.

*Added value of this study:* We provide a comprehensive assessment of the performance of the SPARRA score across a range of population subgroups in several ways. We systematically examined differences in performance using a range of metrics. We identify notable areas of differential performance associated with age, sex, socioeconomic deprivation, ethnicity and residence location (mainland versus island; urban versus rural). We also examined the pattern of errors in prediction across medical causes of emergency admission, finding that, to variable degrees across groups, cardiac and respiratory admissions are more likely to be correctly predicted from electronic health records. Overall, our work provides an atlas of performance measures for SPARRA and partly explains how between-group performance differences arise.

*Implications of all the available evidence:* The precision by which the SPARRA score can predict emergency hospital admissions differs between population subgroups. These differences are largely driven by variation in performance across age and sex, as well as the predictability of different causes of admission. Awareness of these differences is important when making decisions based on the SPARRA score.

## 1 Introduction

The UK’s healthcare system has been repeatedly reported to be under significant pressure due to increasing workload, hospital demand, and the resulting strain on workforce and resources [1, 2]. The COVID-19 pandemic and its aftermath have intensified the challenges faced by human resources in primary care [3]. Consequently, proactive interventions to prevent individuals from experiencing abrupt breakdowns in health have been highlighted as a key priority for modern medical practice [4]. In particular, emergency hospital admissions (EAs), in which urgent in-hospital care is needed for an individual, are a potential target for primary care intervention, as some of these events can be averted through appropriate anticipatory care [5, 6, 7]. However, since total primary care capacity is limited, it is important to optimise the allocation of existing resources [8]. To this end, individual-level prediction of future EAs risk can shape decision-making by helping to identify individuals who may benefit the most from anticipatory primary care intervention [9, 10, 11, 12, 13, 14].

SPARRA (Scottish Patients At Risk of Readmission and Admission) is a risk prediction score calculated by Public Health Scotland (PHS) using routinely collected Scotland-wide electronic health records [15] to estimate, at an individual level, the probability of having an emergency in-patient hospital admission in the subsequent year. To date, three versions of the SPARRA score have been deployed nationally in Scotland, the first one dating back to 2006. SPARRA version 3 (referred to as SPARRAv3) [16] has been in use since 2012. Each month, PHS calculates SPARRAv3 scores for around 80% of Scottish population. A fourth version (SPARRAv4) was recently developed [15] and is expected to be deployed by 2024 on a national scale. SPARRA scores aim to support General Practitioners (GPs) as they plan anticipatory interventions for the patients under their care. At an aggregate level, the scores can also be used to e.g. predict future hospital demand.

Our objective is to study the differential behaviour and accuracy of the SPARRA score across population subgroups (such as urban or rural populations, and residents of more or less deprived geographic areas). We expect that the behaviour of the score may vary across such subgroups due to variable demographic characteristics (age, sex, and deprivation), and potentially due to differential data availability and access to healthcare [17]. Even if the SPARRA score very accurately predicts EA risk in all groups, between-group differences in demographic characteristics may mean that decisions made on the basis of the SPARRA score may have different consequences across different groups, in that if practitioners take action on all patients for whom the SPARRA score exceeds a given threshold, the rates of false positives and false negatives may vary between groups. We consider that without awareness of these differential consequences, primary care practitioners may be unreasonably dissuaded from using SPARRA scores, potentially introducing or exacerbating existing health inequalities (in which Scotland ranks poorly compared to western and central Europe [18, 19]). Hence, it is critical to scrutinise its behaviour across a range of demographic groups, which can also help better understand the epidemiology behind EAs in Scotland.

In this article, we present a retrospective comprehensive evaluation of the performance of SPARRA across a range of groupings defined by age, sex, deprivation, ethnicity, and geographic location (rural versus urban; island versus mainland residency). Our results and code are publicly available on Github and the SPARRAfairness R package. We also provide an online Shiny application for interactive exploration of our results. We intend it to be usable by primary care practitioners and the public to enable informed decision-making and enhance the interpretation of SPARRA scores.

## 2 Methods

### 2.1 Data

Our primary analysis is based on the third version of the score (SPARRAv3), deployed at a national level in Scotland since 2012 [16] (see Supplementary Note S3.2 for details). We repurposed the same retrospective data and inclusion/exclusion criteria used by [15], focusing on a single prediction time cutoff (May 1st, 2016). The data comprise every acute hospital record (EAs, elective admissions, day cases, outpatient attendances, A&E attendances, and records of long-term conditions) and community prescribing activity within the National Health Service (NHS) Scotland up to three years before the time cutoff. We also use age, sex, and mortality records, and long-term condition (LTC) records dating back to January 1981.

For this analysis, the data were extended to include information that is not currently used as input for SPARRA. First, self-reported ethnicity information was obtained by PHS through cross-reference with a range of datasets, including the COVID-19 vaccination programme (see Supplementary Note S3.10). Moreover, PHS used postcode information to derive urban/rural and mainland/island residence status indicators (considered dichotomously as whether an individual lives on the mainland or on any Scottish island). The separation between urban and rural postcodes was based on the Scottish government’s 2016 classification system [20].

### 2.2 Definition of demographic groups

We consider various aspects of the performance of SPARRAv3 over demographically-defined cohorts of patients defined by age, sex (female/male as per current Scottish community health index number), SIMD quintiles (with lower quintiles indicating higher deprivation), ethnicity and indicators for urban/rural and mainland/island residence status. Hereafter, we refer to these as *grouping variables*. For each grouping variable, missing values were excluded when defining the associated groups (rates of missingness are shown in Table 2). For ease of interpretation, we considered only a two-group comparison for age and SIMD. Rather than dichotomising the entire population, we chose groups to highlight the differences between the extremes of these variables (over 65 and under 25 for age; most and least deprived quintiles for SIMD, derived from the top and bottom two deciles). Ethnicity data was also aggregated into two groups (white and non-white) due to the small sample sizes observed for some non-white groups.

Our choice of grouping variables was restricted to those identifiable from data held by PHS, whilst capturing commonly-identified sources of inequality. In particular, we considered groups defined by urban/rural postcodes and by mainland/island postcodes given the potential variation across these groups in environmental and socioeconomic factors (such as employment opportunities) and in access to healthcare [21, 22]. We considered ethnicity as a grouping variable due to its association with differential access to healthcare, socioeconomic and environmental factors, and cultural practices [23, 24, 25]. None of these variables are used as inputs for the SPARRA score. Finally, we included age, sex and deprivation (SIMD), all of which are used as inputs for SPARRA. Deprivation was considered as growing evidence suggests that health inequalities among Scotland’s most and least deprived areas are substantial [26]. Age and sex were selected as risk factors that are known to influence EA susceptibility, disease patterns and outcomes.

### 2.3 Metrics

To deliver a comprehensive evaluation of SPARRA and its performance across different groups, we applied a variety of metrics recently employed across the ML literature, generally in the context of between-group fairness [27, 28, 29]. These comprise measures of predictive discrimination and calibration as well as different types of error rates (see Table 1 and Supplementary Note S3 for more detailed definitions).

**Table 1:**
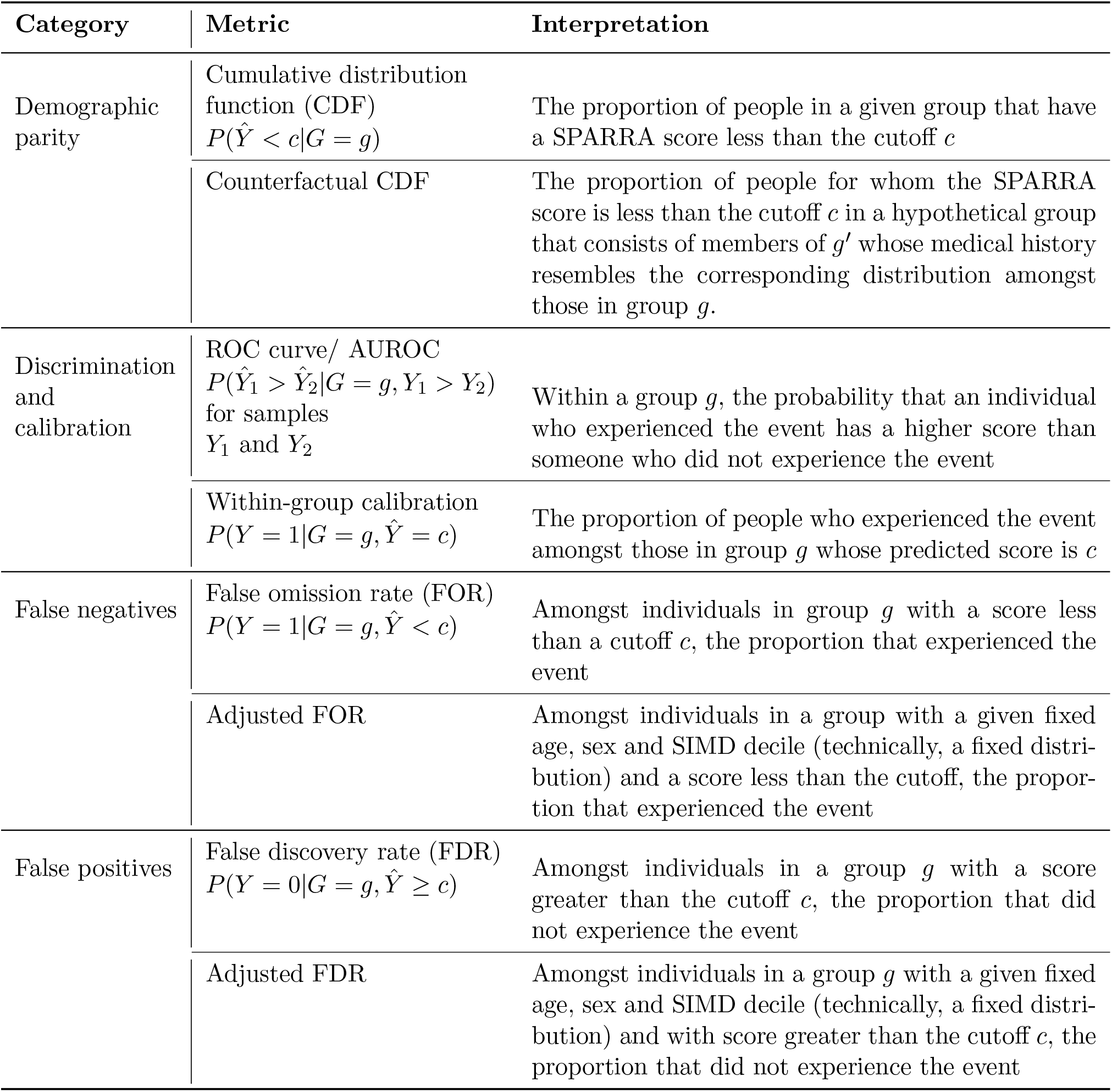
Metrics used to assess score and a brief interpretation, including the quantity we aim to estimate. We compare estimated probabilities across two groups, *G* = *g* and *G* = *g*^*′*^, and a particular score cutoff *c*. For details and formal definitions, see Supplementary Section S3.

| Category | Metric | Interpretation |
| --- | --- | --- |
| Demographic parity | Cumulative distribution function (CDF)<br>$P(\hat{Y} < c \vert G = g)$ | The proportion of people in a given group that have a SPARRA score less than the cutoff $c$ |
| | Counterfactual CDF | The proportion of people for whom the SPARRA score is less than the cutoff $c$ in a hypothetical group that consists of members of $g'$ whose medical history resembles the corresponding distribution amongst those in group $g$. |
| Discrimination and calibration | ROC curve/ AUROC<br>$P(\hat{Y}_1 > \hat{Y}_2 \vert G = g, Y_1 > Y_2)$<br>for samples $Y_1$ and $Y_2$ | Within a group $g$, the probability that an individual who experienced the event has a higher score than someone who did not experience the event |
| | Within-group calibration<br>$P(Y = 1 \vert G = g, \hat{Y} = c)$ | The proportion of people who experienced the event amongst those in group $g$ whose predicted score is $c$ |
| False negatives | False omission rate (FOR)<br>$P(Y = 1 \vert G = g, \hat{Y} < c)$ | Amongst individuals in group $g$ with a score less than a cutoff $c$, the proportion that experienced the event |
|  | Adjusted FOR | Amongst individuals in a group with a given fixed age, sex and SIMD decile (technically, a fixed distribution) and a score less than the cutoff, the proportion that experienced the event |
| False positives | False discovery rate (FDR)<br>$P(Y = 0 \vert G = g, \hat{Y} \geq c)$ | Amongst individuals in a group $g$ with a score greater than the cutoff $c$, the proportion that did not experience the event |
| | Adjusted FDR | Amongst individuals in a group $g$ with a given fixed age, sex and SIMD decile (technically, a fixed distribution) and with score greater than the cutoff $c$, the proportion that did not experience the event |

**Table 2:** Descriptive statistics stratified across groups (in columns). **N** was rounded to the nearest thousand. All numbers are percentages unless indicated otherwise. *Italicised* groups are not specifically analysed. ‘Most’: in the most deprived quintile; ‘Least’: in the least deprived quintile; ‘Miss.’: missing. There were no missing values for age, sex, or deprivation.

|  | *All* | **Sex** |  | **Age** |  |  | **Deprivation** |  |  |
| --- | --- | --- | --- | --- | --- | --- | --- | --- | --- |
| | | M | F | $\geq 65$ | $\leq 25$ | *26-64* | Most | Least | *Other* |
| **N (thousand)** | *4286* | 1946 | 2340 | 884 | 1100 | *2302* | 923 | 803 | *2560* |
| **Sex and age** |  |  |  |  |  |  |  |  |  |
| Male | *45.4* | 100 | 0 | 44.1 | 47.5 | *44.9* | 46.2 | 44.8 | *45.3* |
| Age (years, mean) | *43.3* | 42.7 | 43.7 | 75.5 | 12.1 | *45.8* | 40.4 | 44.1 | *44* |
| Age (years, SD) | *23.8* | 23.9 | 23.7 | 7.22 | 7.51 | *11.7* | 23.2 | 24.1 | *23.7* |
| **Deprivation** |  |  |  |  |  |  |  |  |  |
| Most deprived | *21.5* | 21.9 | 21.2 | 17.1 | 23.8 | *22.2* | 100 | 0 | *0* |
| Least deprived | *18.7* | 18.5 | 18.9 | 20.3 | 18.9 | *18.1* | 0 | 100 | *0* |
| **Ethnicity** |  |  |  |  |  |  |  |  |  |
| White (W) | *66.8* | 65.1 | 68.1 | 54.5 | 69.7 | *70.1* | 70.3 | 64.6 | *66.2* |
| Nonwhite (NW) | *21.8* | 23 | 20.8 | 13.9 | 24.9 | *23.3* | 18.3 | 23.9 | *22.3* |
| **Postcode** |  |  |  |  |  |  |  |  |  |
| Urban (U) | *83.3* | 83 | 83.6 | 80.3 | 85 | *83.7* | 96.5 | 91.9 | *75.9* |
| Rural (R) | *16.6* | 16.9 | 16.4 | 19.7 | 15 | *16.2* | 3.47 | 8.07 | *24* |
| Mainland (ML) | *98.1* | 98 | 98.1 | 97.6 | 98.4 | *98.1* | 99.7 | 99.7 | *96.9* |
| Island (IL) | *1.86* | 1.91 | 1.82 | 2.38 | 1.57 | *1.8* | 0.184 | 0.222 | *2.98* |

Table 2: Descriptive statistics stratified across groups (in columns).
|  | *All* | **Ethnicity** |  |  | **Urban/rural** |  |  | **Mainland/Island** |  |
| --- | --- | --- | --- | --- | --- | --- | --- | --- | --- |
|  |  | W | NW | *Miss.* | U | R | *Miss.* | ML | IL |
| **N (thousand)** | *4286* | 2862 | 933 | *491* | 3571 | 713 | *3* | 4204 | 80 |
| **Sex and age** |  |  |  |  |  |  |  |  |  |
| Male | *45.4* | 44.3 | 47.9 | *47.1* | 45.2 | 46.2 | *50.9* | 45.4 | 46.7 |
| Age (years, mean) | *43.3* | 41.4 | 39.3 | *61.8* | 42.7 | 46 | *42* | 43.2 | 47.3 |
| Age (years, SD) | *23.8* | 22.6 | 22.1 | *31.7* | 23.7 | 23.9 | *22.2* | 23.7 | 24 |
| **Deprivation** |  |  |  |  |  |  |  |  |  |
| Most deprived | *21.5* | 22.7 | 18.2 | *21.3* | 24.9 | 4.49 | *22.5* | 21.9 | 2.13 |
| Least deprived | *18.7* | 18.1 | 20.6 | *18.7* | 20.7 | 9.09 | *12.4* | 19 | 2.23 |
| **Ethnicity** |  |  |  |  |  |  |  |  |  |
| White (W) | *66.8* | 100 | 0 | *0* | 67.2 | 64.4 | *62.2* | 66.9 | 58.3 |
| Nonwhite (NW) | *21.8* | 0 | 100 | *0* | 21.3 | 24 | *27.1* | 21.6 | 29 |
| **Postcode** |  |  |  |  |  |  |  |  |  |
| Urban (U) | *83.3* | 83.9 | 81.6 | *83.2* | 100 | 0 | *0* | 84.5 | 26 |
| Rural (R) | *16.6* | 16 | 18.3 | *16.8* | 0 | 100 | *0* | 15.5 | 74 |
| Mainland (ML) | *98.1* | 98.3 | 97.4 | *97.9* | 99.4 | 91.7 | *0* | 100 | 0 |
| Island (IL) | *1.86* | 1.63 | 2.48 | *2.06* | 0.58 | 8.29 | *0* | 0 | 100 |

We will use *Y ∈ {*0, 1*}* to denote the observed outcome for an individual, where *Y* = 1 means an EA or death occurs within one year from the prediction time cutoff. We use *G* to denote the group under consideration to which the individual belongs. We denote the SPARRAv3 score for the individual as *Ŷ*. We consider a hypothetical decision rule that binarises those predictions such that the decision is to predict *Y* = 1 for an individual if and only if *Ŷ> c* and consider the consequences of varying *c* across the interval [1, 99]%.

We first assess ‘demographic parity’ by comparing the cumulative distribution of scores *Ŷ* in the grouped populations, i.e., *P* (*Ŷ< c*|*G* = *g*). We then compare the distribution of scores under a ‘counterfactual’ setting in which we substitute a grouping value for another (e.g. rural instead of urban), while the underlying distribution of hospital activity data and prescriptions is held constant [30, 31]. This isolates the effects of the grouping variable on the score to only those mediated through either the direct effect of the group or through differences in distributions of age, sex and SIMD.

We evaluated group-level predictive performance using receiver-operator characteristic curve (ROC) and calibration curves. The area under the ROC (AUROC) quantifies the ability of the model to rank individuals accurately based on predicted risk; we thus evaluate the model’s ability to discriminate between predicted admissions versus non-admissions for all groups under consideration. Note that predicted risk can inaccurately represent observed risk even if the model attains good discrimination [32]. Instead, calibration assesses the agreement between the observed and predicted number of events [33]. A risk score that is well-calibrated for group *g* should have *P* (*Y* = 1|*G* = *g, Ŷ* = *c*) *≈ c* for most *c*; that is, amongst individuals with *Ŷ* = *c*, a proportion of around *c* have *Y* = 1. We assessed calibration visually by directly plotting calibration curves (sometimes called reliability diagrams [34]) in each group.

We then assess the rates of false-negative and false-positive errors [35] by group based on the hypothetical decision rule. A false negative means a setting in which the SPARRA score predicted that an individual would not have an event (i.e., *Ŷ< c*, for a given cutoff value *c*), but that individual did have an event (i.e. *Y* = 1 is observed). Correspondingly, a false positive means a setting for which *Ŷ≥ c* but *Y* = 0. Within a group *g*, the probability of observing a false negative is *P* (*Y* = 1|*G* = *g, Ŷ < c*), and the probability of observing a false positive is *P* (*Y* = 0|*G* = *g, Ŷ≥ c*), which we term false omission rate (FOR) and false discovery rate (FDR) respectively. We considered both direct estimates of FOR and FDR and adjusted estimates to remove mediating effects of age, sex, and SIMD. These adjusted metrics may be considered as counterfactual values after substituting one group with another (Supplementary Section S3.5.2). Note that it is possible for a risk score to be perfectly calibrated in two groups *G* = *g* and *G* = *g*^*′*^ and still have differing FOR or FDR between groups. Moreover, in our context, false negatives are of somewhat greater concern than false positives, as they represent individuals who potentially missed out on treatment. High false positive error rates may trigger unnecessary interventions and excessive costs to the healthcare system.

Finally, to better understand the nature of false negatives, we assessed the extent to which various causes of admission (using the first letter of the ICD10 code recorded as a primary admission diagnosis, Supplementary Table 3) could be predicted in each group. For this purpose, we considered the population of individuals with SPARRA scores less than 10% (*Ŷ <* 0.1) which was a false negative; that is, who subsequently had an EA or died within 1 year (*Y* = 1). For individuals who died without having an EA first, we considered the ICD10 code associated with their primary cause of death. Such individuals essentially constitute an extreme example, in which an EA or death occurred against expectation.

## 3 Results

### 3.1 Data summary

Demographic details for the individuals present in our data are shown in Table 2. Our data comprised slightly more than 50% females and slightly more than 20% individuals in the most-deprived SIMD quintile (and slightly fewer than 20% in the least-deprived quintile).

**Table 3:**
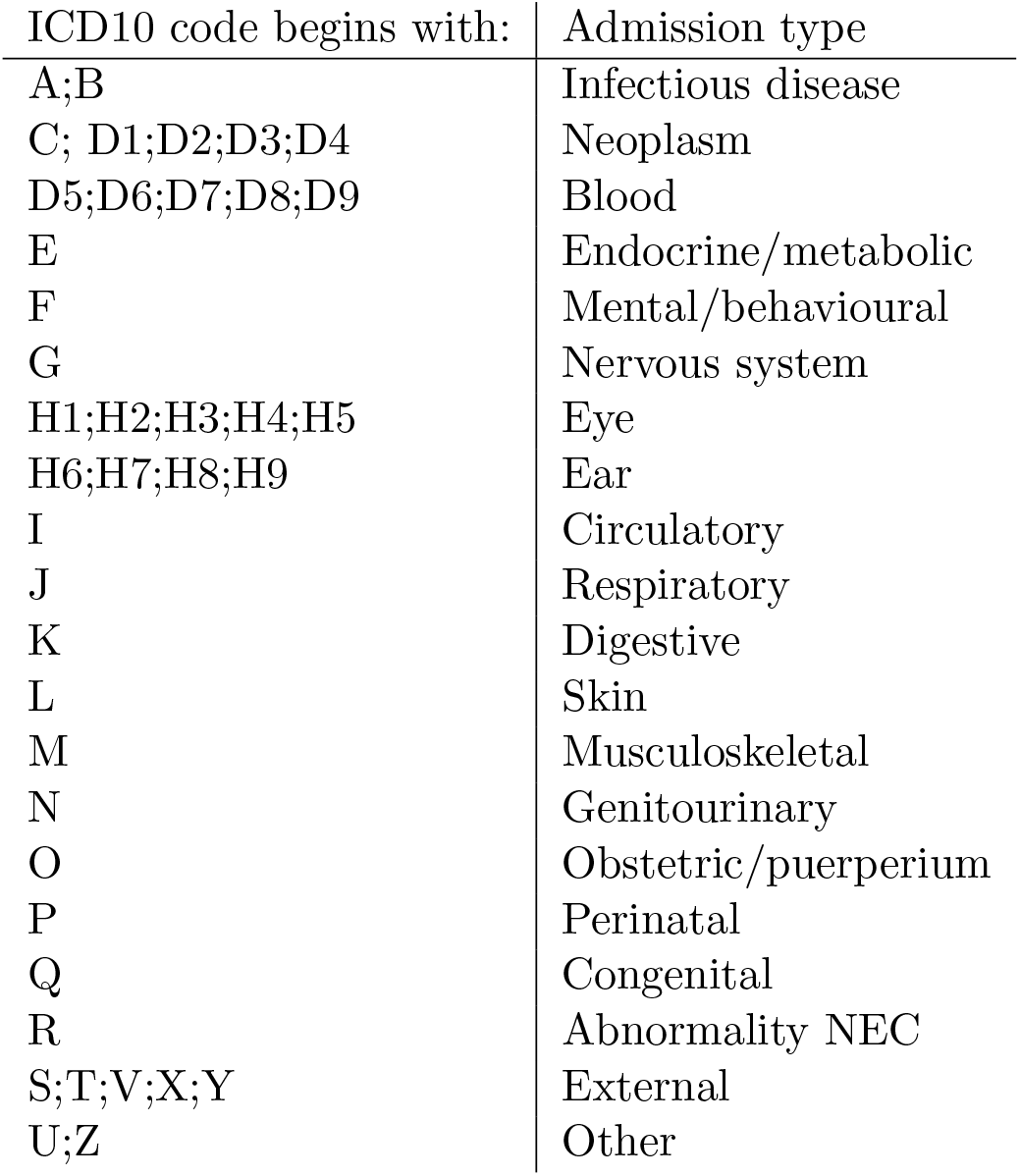
Definition of different admission types.

| ICD10 code begins with: | Admission type |
| --- | --- |
| A;B | Infectious disease |
| C; D1;D2;D3;D4 | Neoplasm |
| D5;D6;D7;D8;D9 | Blood |
| E | Endocrine/metabolic |
| F | Mental/behavioural |
| G | Nervous system |
| H1;H2;H3;H4;H5 | Eye |
| H6;H7;H8;H9 | Ear |
| I | Circulatory |
| J | Respiratory |
| K | Digestive |
| L | Skin |
| M | Musculoskeletal |
| N | Genitourinary |
| O | Obstetric/puerperium |
| P | Perinatal |
| Q | Congenital |
| R | Abnormality NEC |
| S;T;V;X;Y | External |
| U;Z | Other |

In total, 66.8% of the cohort had a recorded white ethnicity and 21.8% non-white. A substantial proportion (11.5%) of the study population had no ethnicity reported, and the absence of ethnicity data was non-random. Individuals with missing ethnicity were generally older, with higher variability of ages, potentially due to differential uptake of the COVID-19 vaccine. However, they did not include disproportionate numbers of members of the most- or least-deprived quintiles or urban or mainland postcodes (Table 2).

As expected, the majority of the cohort was urban (83.3%) rather than rural, and a majority lived on the mainland (98.1%) rather than on islands. For 2,837 (*<* 0.07%) included samples, it was not possible to resolve the postcode to an urban-rural status or a mainland-island status. The cohort of samples for which the individual was missing a postcode had a higher proportion of males, but otherwise reasonably typical of the general cohort.

### 3.2 Score distribution and performance

We directly analysed the distribution of SPARRA scores in each group, using cumulative distributions (also called demographic parity) and a counterfactual substitution of the alternative group. Despite sex being an input for SPARRA, there are no substantial differences in the distribution of scores for males and females (Supplementary Figure 4b). A similar pattern was observed when comparing groups defined by urban/rural (Supplementary Figure 4e) or mainland/island (Supplementary Figure 7f) residence status. The largest difference was observed when comparing different age groups: as can be expected, individuals over 65 had much higher scores than those under 25 (Supplementary Figure 4a). The difference in score distributions between these age groups was smaller when we used a counterfactual comparison (Supplementary Figure 5a), indicating that a large part of the difference is due to between-group differences data on hospital activity and prescriptions, with the residual difference due to direct effects of age on the score (and potentially due to effects mediated through sex or SIMD distributions).

Whilst we initially observed a difference in the distribution of the scores between nonwhite and white individuals (Figure 1a), the difference largely disappeared in the counterfactual comparison (Figure 1c). Individuals in the most deprived quintile tend to have higher scores than those in the least deprived quintile (Figure 1b). This effect was somewhat reduced in the counterfactual comparison, but still present (Figure 1d). The difference in counterfactual score distributions can only be due to different distributions of age and sex in SIMD quintiles and by direct effects of SIMD on score (SIMD is an input for SPARRA). However, it is unlikely to be solely due to the former, as individuals in the most deprived quintile were of *lower* average age (whose EA risk tends to be lower), and there was little difference in sex distribution between the most and least deprived quantiles (Table 2). This indicates that the differences in counterfactual score distribution are due to the direct effects of SIMD on the risk score.

**Figure 1:**
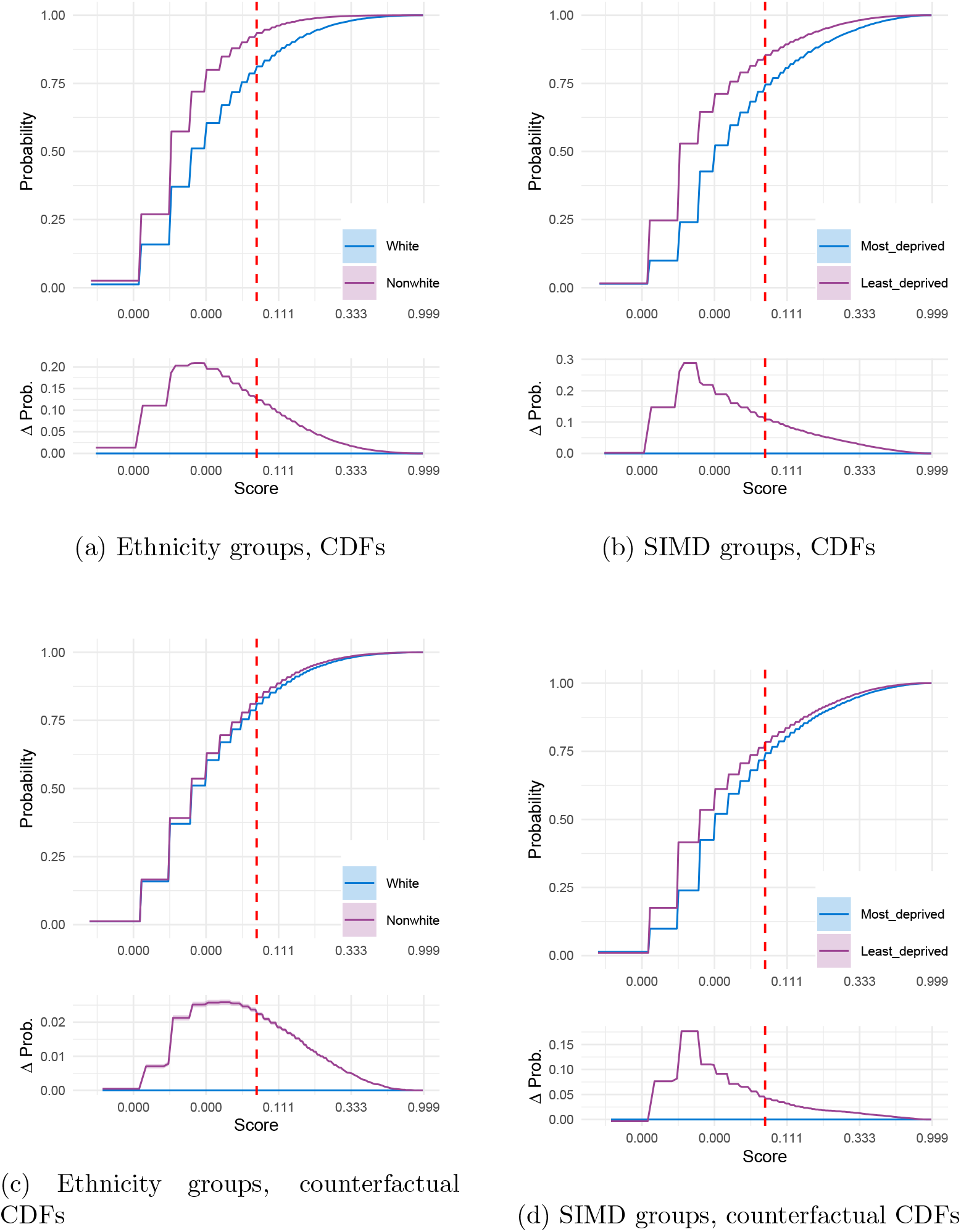
Demographic parity, measured by CDF of raw scores and ‘counterfactual’ scores in groups defined by ethnicity and SIMD. Counterfactual scores remove group effects on the score, which are due to different distributions of hospital activity and prescription data between groups, essentially isolating group effects to those mediated through age, sex, and SIMD. Lower sub-panels on each panel show the difference between curves. In all panels, the x-axis is in a logarithmic scale. Coloured bands show 95% confidence bands, though these are often narrow. A right-shifted distribution indicates generally higher scores.

Discrimination (measured by AUROC) was generally stronger in individuals over 65 than under 25. Both white and nonwhite subgroups had poorer discrimination than the overall cohort. SPARRAv3 was generally well-calibrated across all groups. See Supplementary Section S3.9 for details.

### 3.3 False negatives and false positives

Using our hypothetical decision rule (Section 2.3), an EA event is predicted if the estimated SPARRA score (*Ŷ*) is greater or equal than a score cutoff *c*. In most comparisons, we observed differences in the false positive rates between the associated subgroups, regardless of the choice of *c*. False positive rates were higher in individuals under 25 (Supplementary Figure 10a), the least deprived subgroup (Supplementary Figure 11c), nonwhite individuals (Supplementary Figure 10d), and for individuals with rural (Supplementary Figure 10e) and island postcodes (Supplementary Figure 10f) than the corresponding groups. Females and males had similar false positive rates, with females slightly higher at low thresholds (Supplementary Figure 10b). All differences were virtually identical after adjusting for age, sex, and SIMD, indicating that most differences were not due to these variables (Supplementary Figure 11).

False negative rates substantially differed between all pairs of groups, across all values of *c* (Supplementary Figures 8). For example, individuals over 65 had generally higher false negative rates than those under 25 (Supplementary Figures 8a). When considering subgroups defined by the residence postcode, we observed higher false negative rates in urban versus rural residents (Figure 2a) and for those with a mainland versus island residence (Figure 2b). These differences largely remained after adjusting for age, sex, and SIMD (Supplementary Figure 9). After adjustment, however, the gap between urban-rural groups decreased (Figure 2c), while it increased between mainland and island groups (Figure 2d). This indicated that while urban-rural differences in false negative rates were largely driven by between group differences in distributions of age, sex, and SIMD, the differences in distribution between mainland and island groups tended to cause a reduction in the difference in false-negative rate.

**Figure 2:**
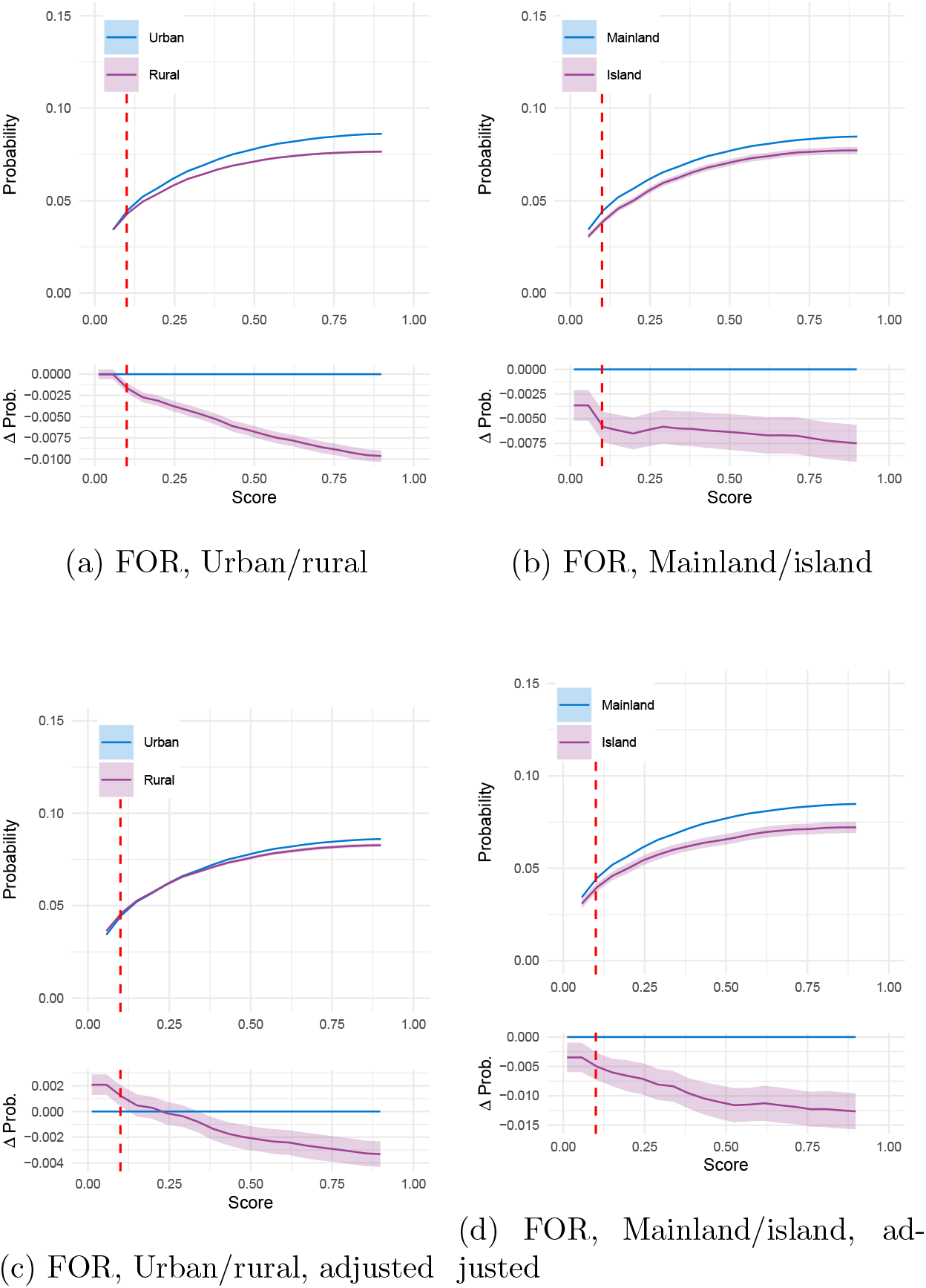
FOR at a range of cutoffs for urban/rural groups (panels 2a, 2c) and mainland/island groups (panels 2b, 2c). Lower panels show FOR adjusted to remove the effect of age, sex, and SIMD. Within each panel, the lower subpanel shows the difference between the two curves on the upper subpanel. Coloured bands indicate 95% pointwise confidence intervals, which are very narrow for some groups.

#### 3.3.1 Decomposition of false negatives

We explored the distribution of admission types within false negatives by considering the cohort of those with a SPARRA score less than 10% (*Ŷ <* 0.1) who subsequently experienced an EA or died within one year. To identify types of admission that are over- or under-represented within this cohort, we compared the frequencies of admission types in this cohort against the frequencies of admission types across all EA (regardless of the outcome predicted by SPARRA). Identifying such admission types is helpful in interpreting the scores, in that scores may be less reassuring against the risk of certain types of admissions than others.

For the overall cohort (Supplementary Figure 12), among all admissions and deaths, the most common recorded reason was due to external causes which in most cases we would not expect to be able to predict (ICD-10 codes S,T,V,X,Y; including accidents, intentional self-harm, assault and medical and surgical complications). Admissions were also frequently listed as ‘abnormality NEC’ (not elsewhere classified; essentially missing). Digestive, respiratory, circulatory and infectious causes of admission were common, whilst neoplastic, blood, ear, eye, and mental/behavioural admissions were rare. As expected, some admission types exhibited sex-specific patterns (e.g. obstetric admissions only recorded for females).

We then calculated the distribution of admission types restricted to individuals in each of the groups considered in this study. External causes of admission were markedly over-represented in subjects with *Ŷ <* 0.1 within several groups, including males (Figure 3a), individuals under 25 (Supplementary Figure 14c), individuals of white ethnicity (Supplementary Figure 14a), those in the most-deprived quintiles (Supplementary Figure 13a). The pattern was still present but to a much lesser extent in some groups, e.g. females (Figure 3b). Generally, respiratory admissions were under-represented amongst those with *Ŷ <* 0.1 within most groups, particularly in individuals over 65 (Supplementary Figure 14d). Circulatory admissions were over-represented in over-65 individuals with *Ŷ <* 0.1, but under-represented in the general population, indicating that they are relatively difficult to predict in over-65s, but relatively easy to predict in the general population (Supplementary Figure 12). Digestive causes of admission were generally disproportionately difficult to predict.

**Figure 3:**
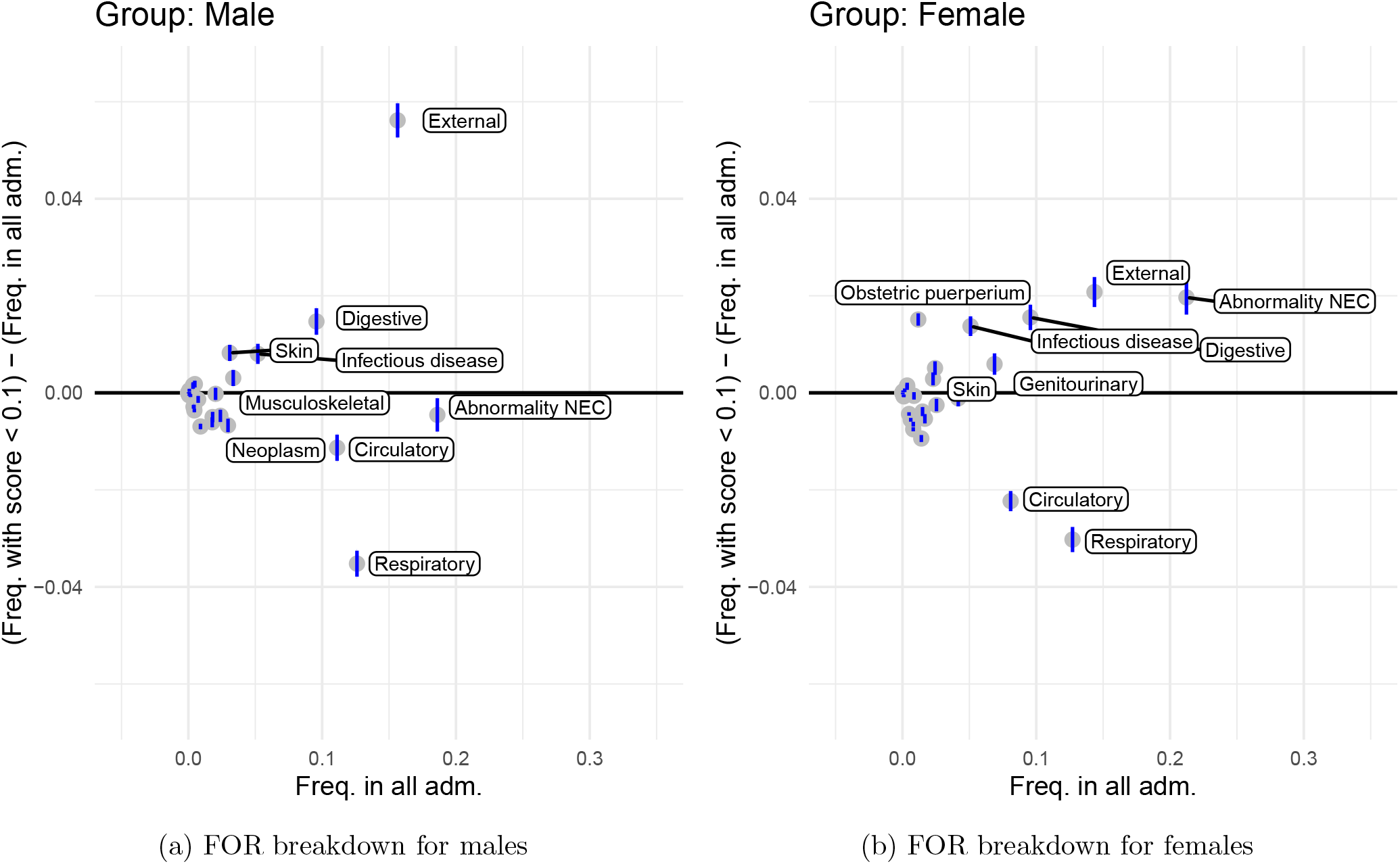
Decomposition of false negatives within males and females. Plots consider the proportion of each admission type amongst all admissions in a group (*A*) and the proportion of each admission type amongst admissions in the group with SPARRA score *<* 10% (*B*) and plots show (*A*) against (*B*) − (*A*). Points above the line *y* = 0 correspond to admissions which are disproportionately poorly identified by SPARRA score (that is, for which *Ŷ <* 0.1). Blue vertical lines show pointwise 95% confidence intervals. Distinctive points are labelled.

**Figure 4:**
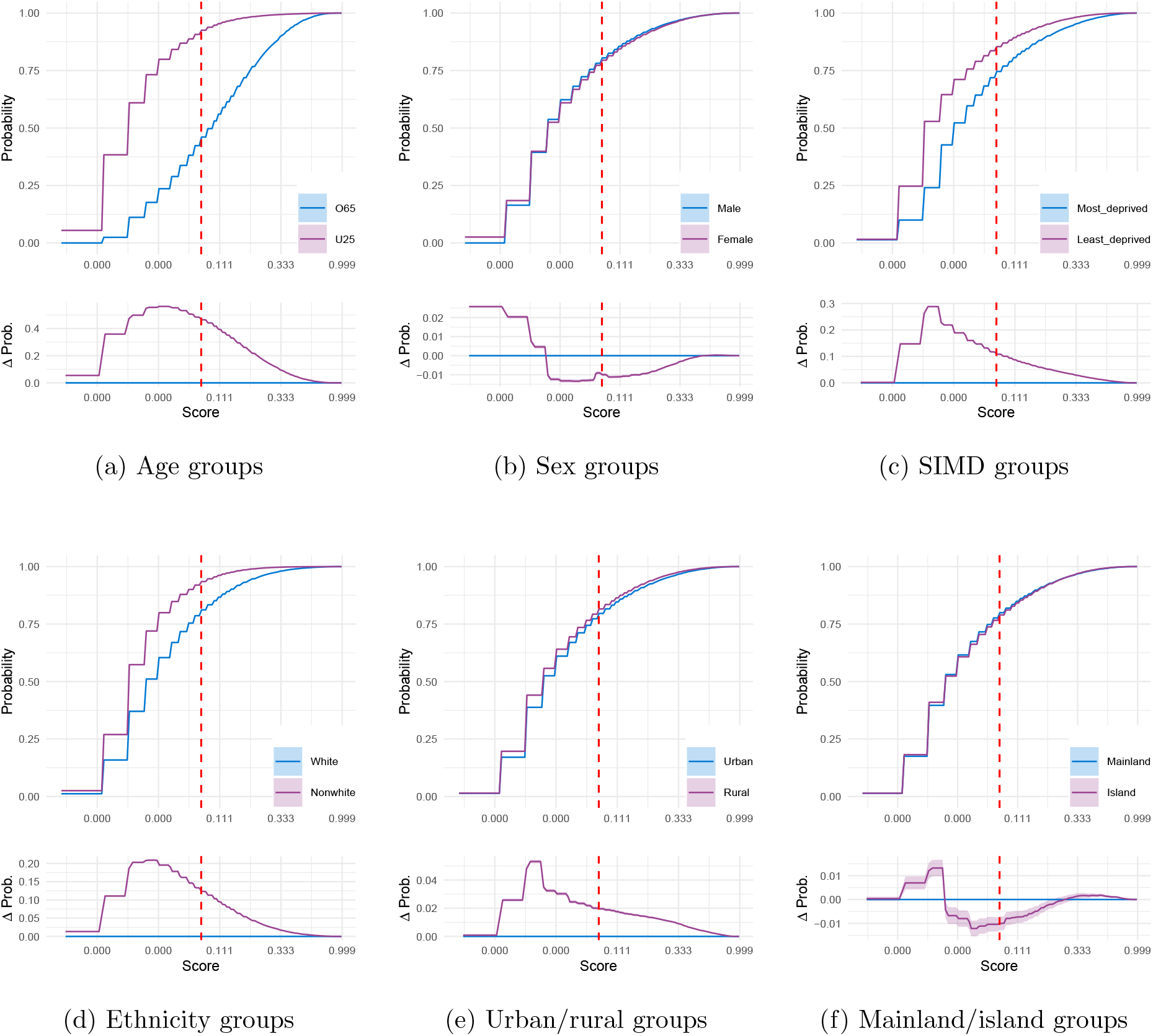
Empirical cumulative distribution of scores (log scaled) in each group, also called demographic parity. Lower panels on each panel show difference between curves. Coloured bands show pointwise 95% confidence intervals. Vertical red dashed lines identify a score of 10%. Figures for age and SIMD are replicated from the main text.

**Figure 5:**
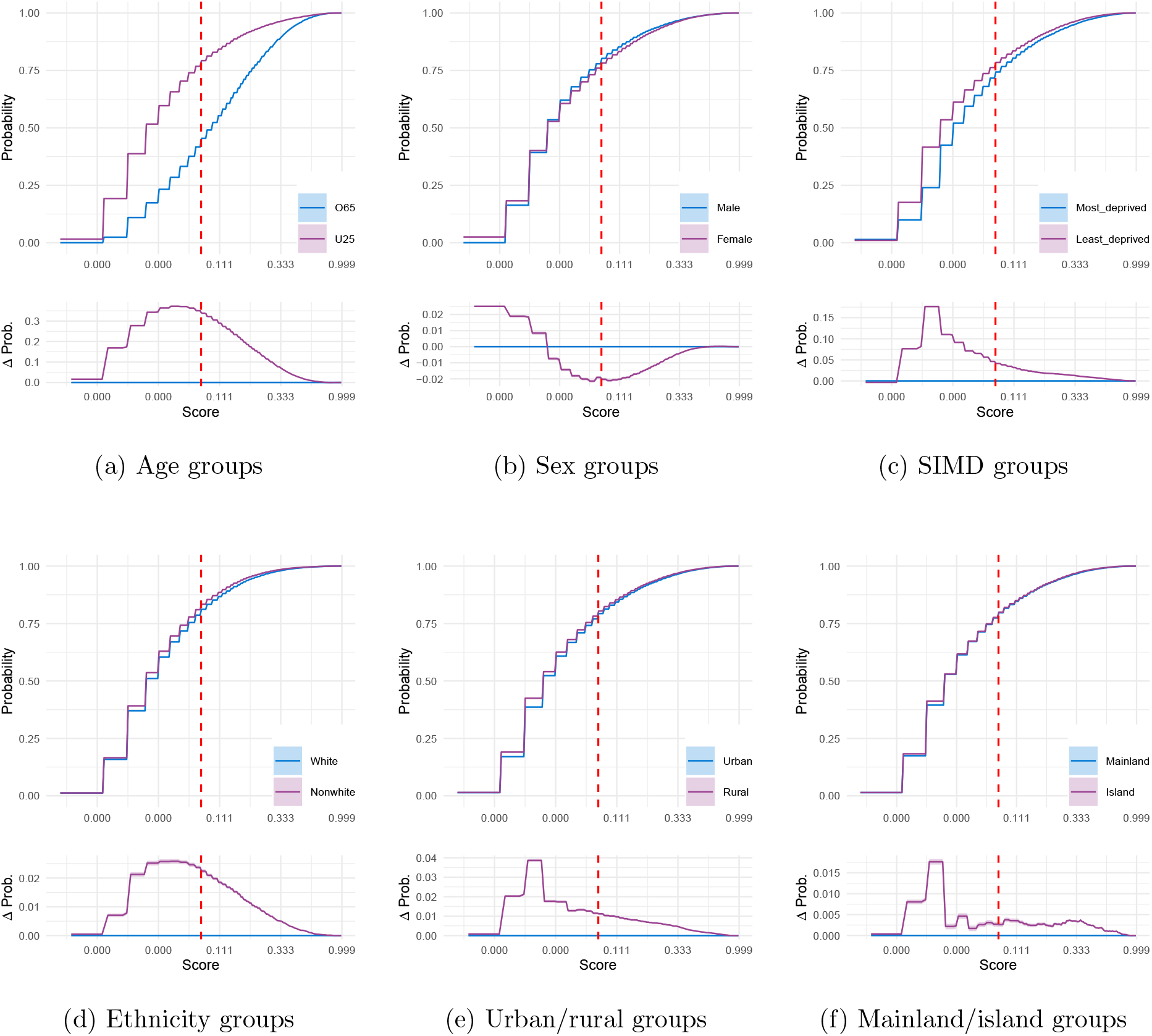
Cumulative distribution of ‘counterfactual’ scores (log scaled) in each group: essentially isolating the effect of group to that mediated through age, sex, and SIMD. Lower panels on each panel show difference between curves. Coloured bands show pointwise 95% confidence intervals. Vertical red dashed lines identify a score of 10%.

**Figure 6:**
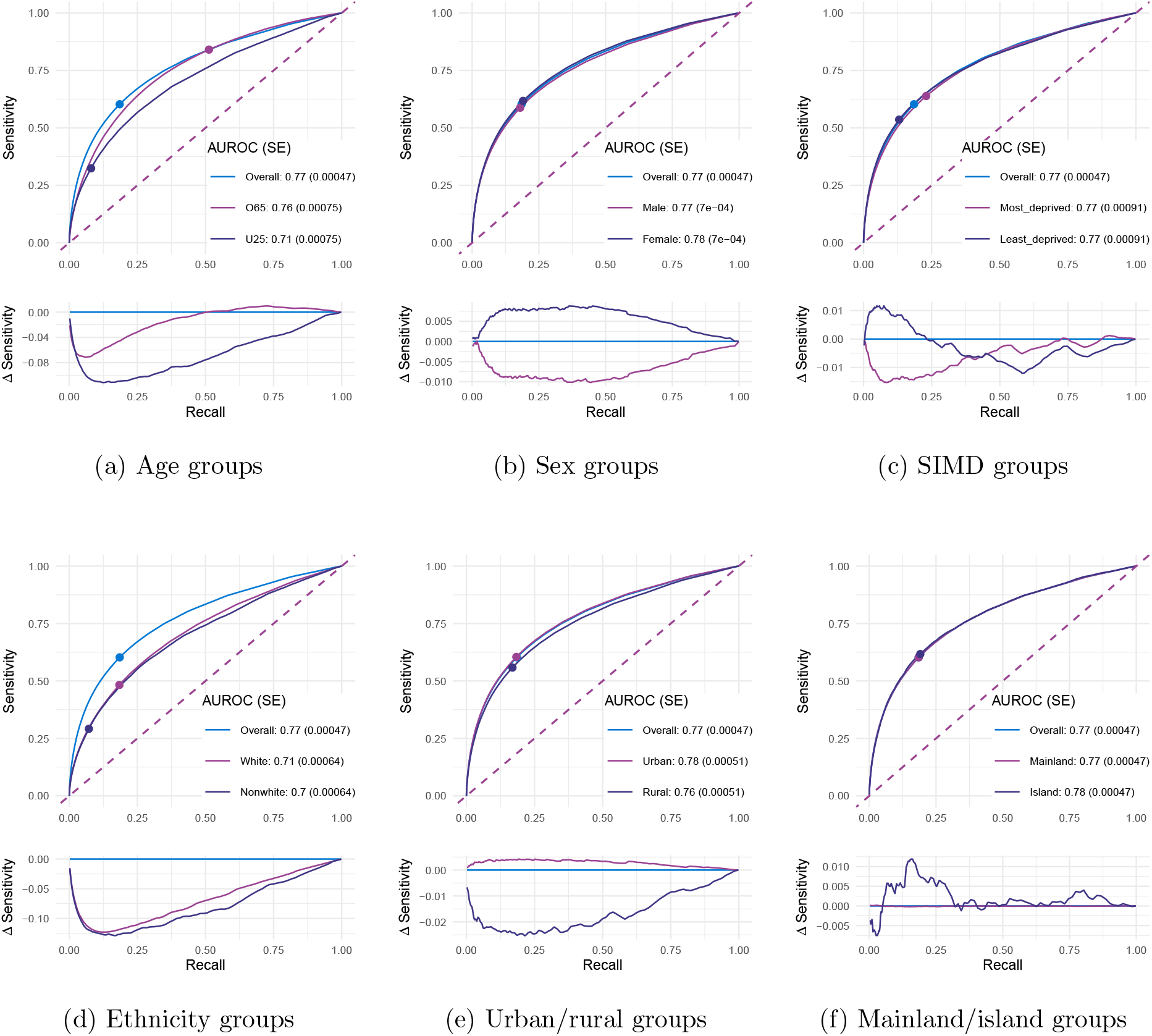
Performance by group assessed using ROC curves (discrimination). Legends show AUROC, and associated standard errors. Lower panels on each panel show difference between curves. Points on figures identify a score of 10%.

**Figure 7:**
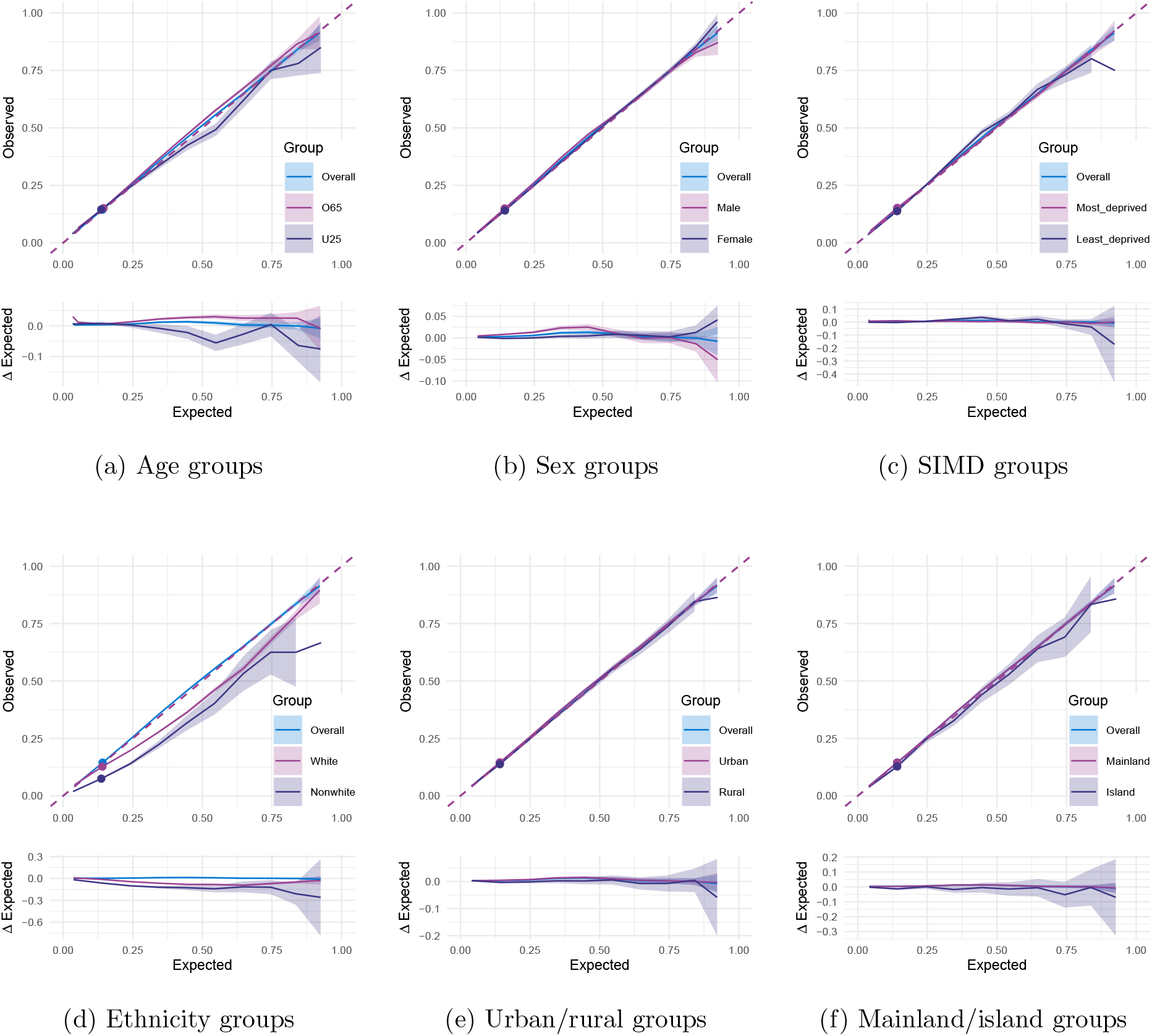
Performance by group assessed using calibration curves. Lower panels on each panel show difference between curves relative to whole group. Coloured bands indicate 95% pointwise confidence intervals. Points on figures identify a score of 10%.

**Figure 8:**
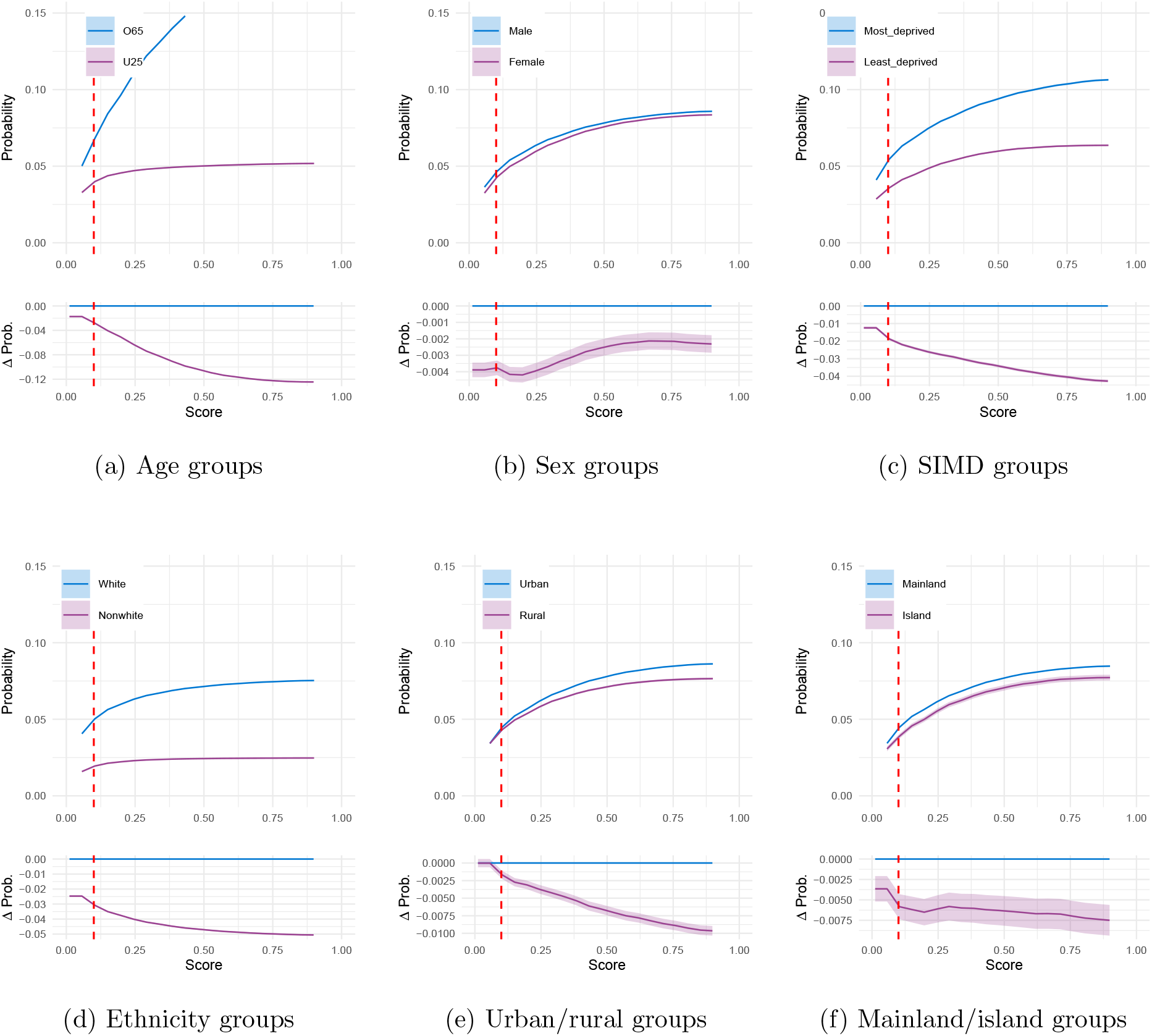
False negative rates by group assessed using FOR (unadjusted); that is, *P* (*Y* = 1|*G* = *g, Ŷ < c*) for cutoff *c* and group *g*. Lower panels on each panel show difference between curves. Coloured bands show pointwise 95% confidence intervals. Vertical red dashed lines identify a score of 10%.

**Figure 9:**
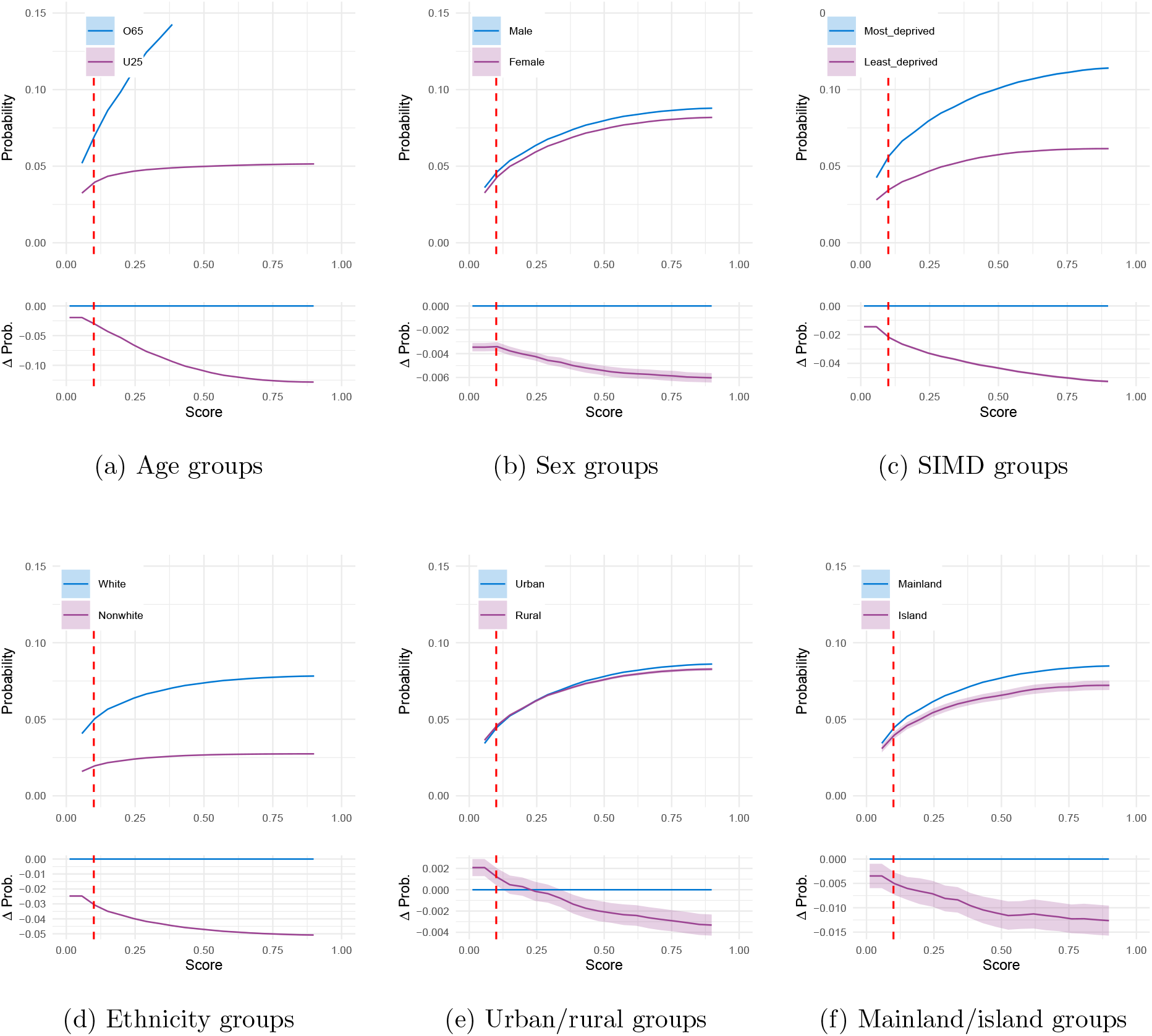
False negative rates by group adjusted for effect of age, sex, and SIMD, thus removing effects mediated by these. Lower panels on each panel show difference between curves. Coloured bands show pointwise 95% confidence intervals. Vertical red dashed lines identify a score of 10%.

**Figure 10:**
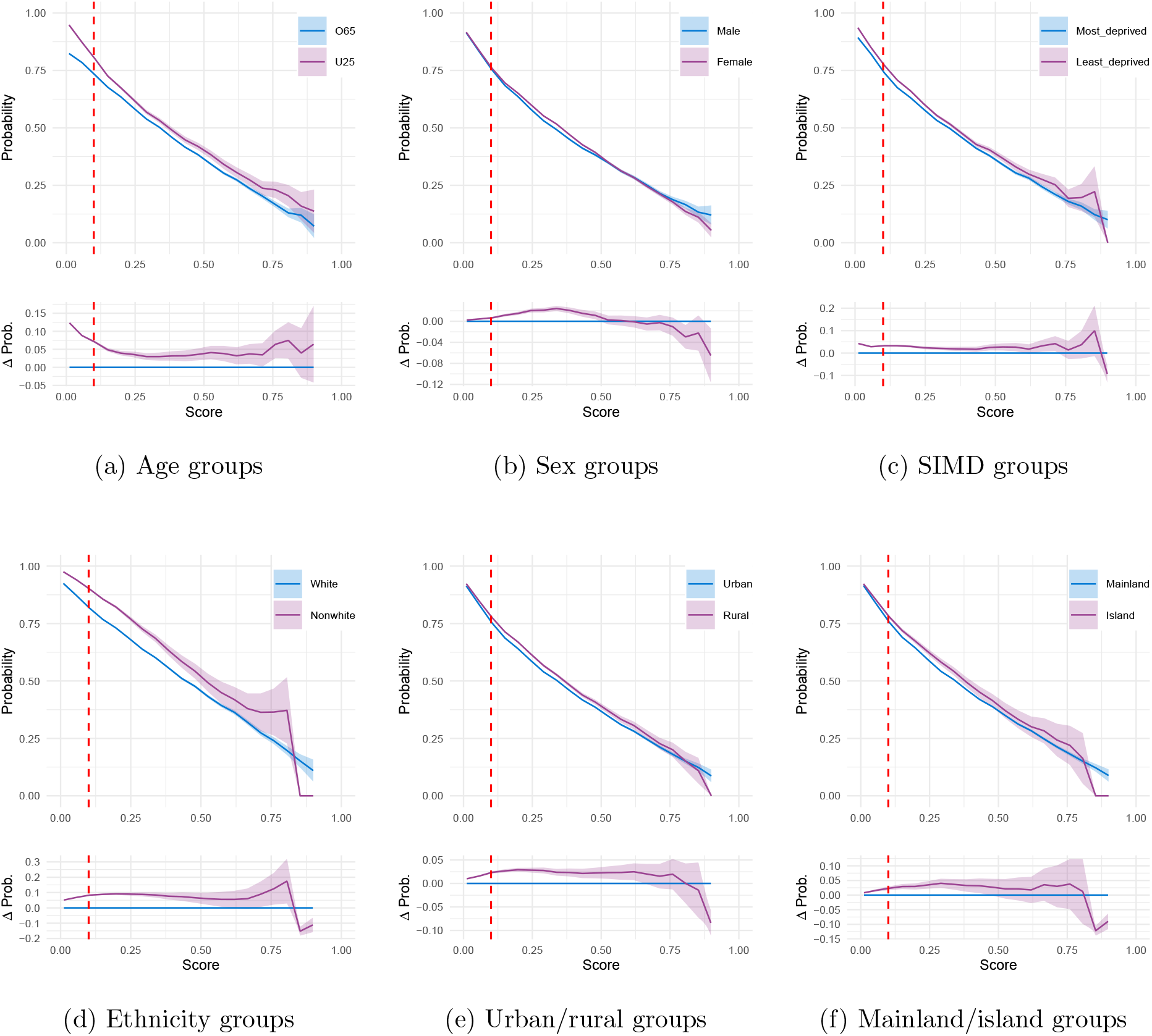
False positive rates by group assessed using FDRP (unadjusted); that is, *P* (*Y* = 0|*G* = *g, Ŷ ≥ c*) for cutoff *c* and group *g*. Lower panels on each panel show difference between curves. Coloured bands show pointwise 95% confidence intervals. Vertical red dashed lines identify a score of 10%.

**Figure 11:**
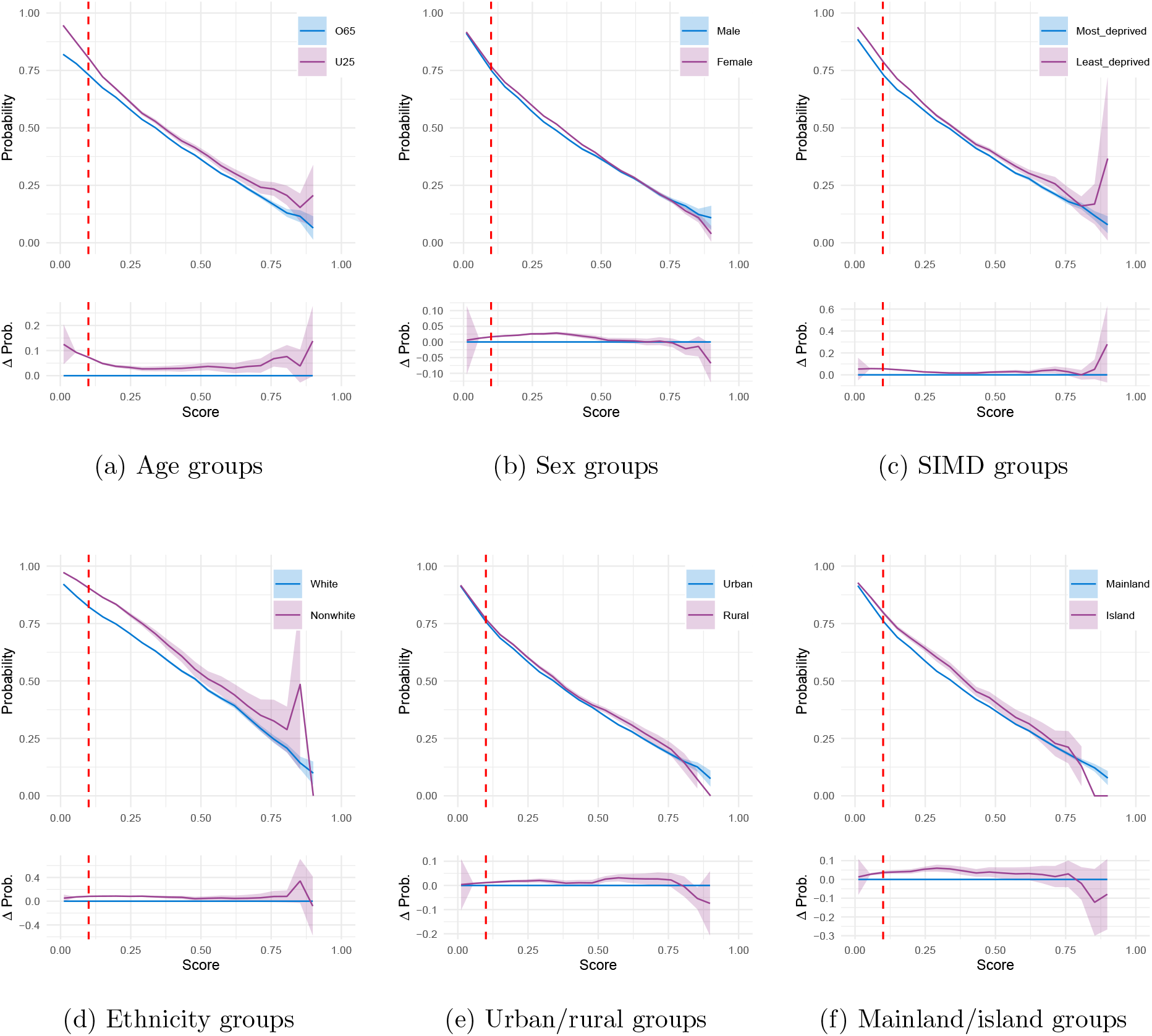
False positive rates by group adjusted for effect of age, sex, and SIMD, thus removing effects mediated by these. Lower panels on each panel show difference between curves. Coloured bands show pointwise 95% confidence intervals. Vertical red dashed lines identify a score of 10%.

**Figure 12:**
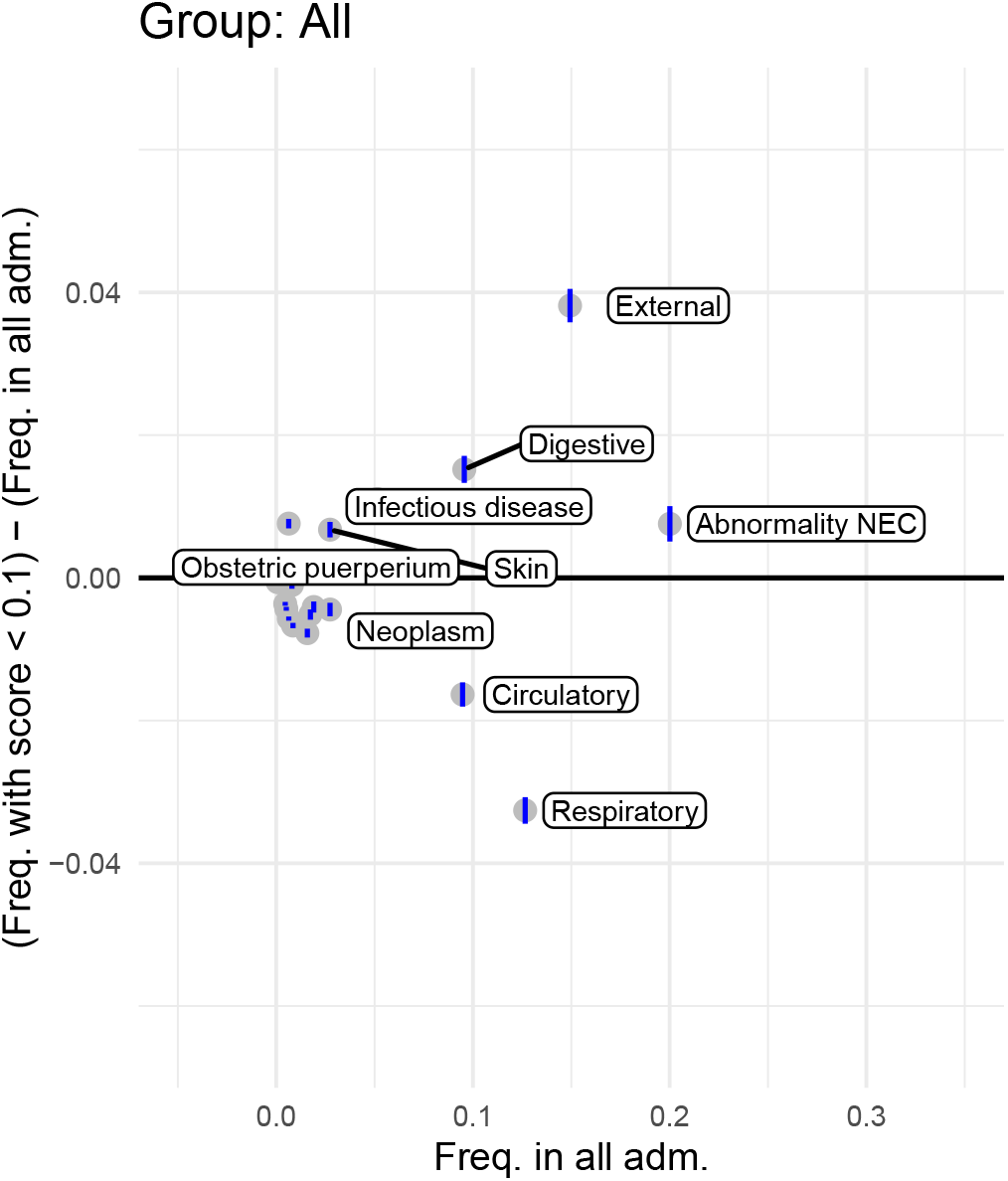
Decomposition of false negatives across all samples. Plots consider the proportion of each admission type amongst all admissions in a group (*A*) and the proportion of each admission type amongst admissions in the group with SPARRA score *<* 10% (*B*) and plots show (*A*) against (*B*) − (*A*). Points above the line *y* = 0 correspond to admissions which are disproportionately poorly identified by SPARRA score (strictly the criterion *Ŷ <* 0.1)., Blue vertical lines show pointwise 95% confidence intervals. Upper plots show the proportion of (unpredicted) admissions due to each cause; lower plots show the proportion of deaths due to each cause. Distinctive points are labelled.

**Figure 13:**
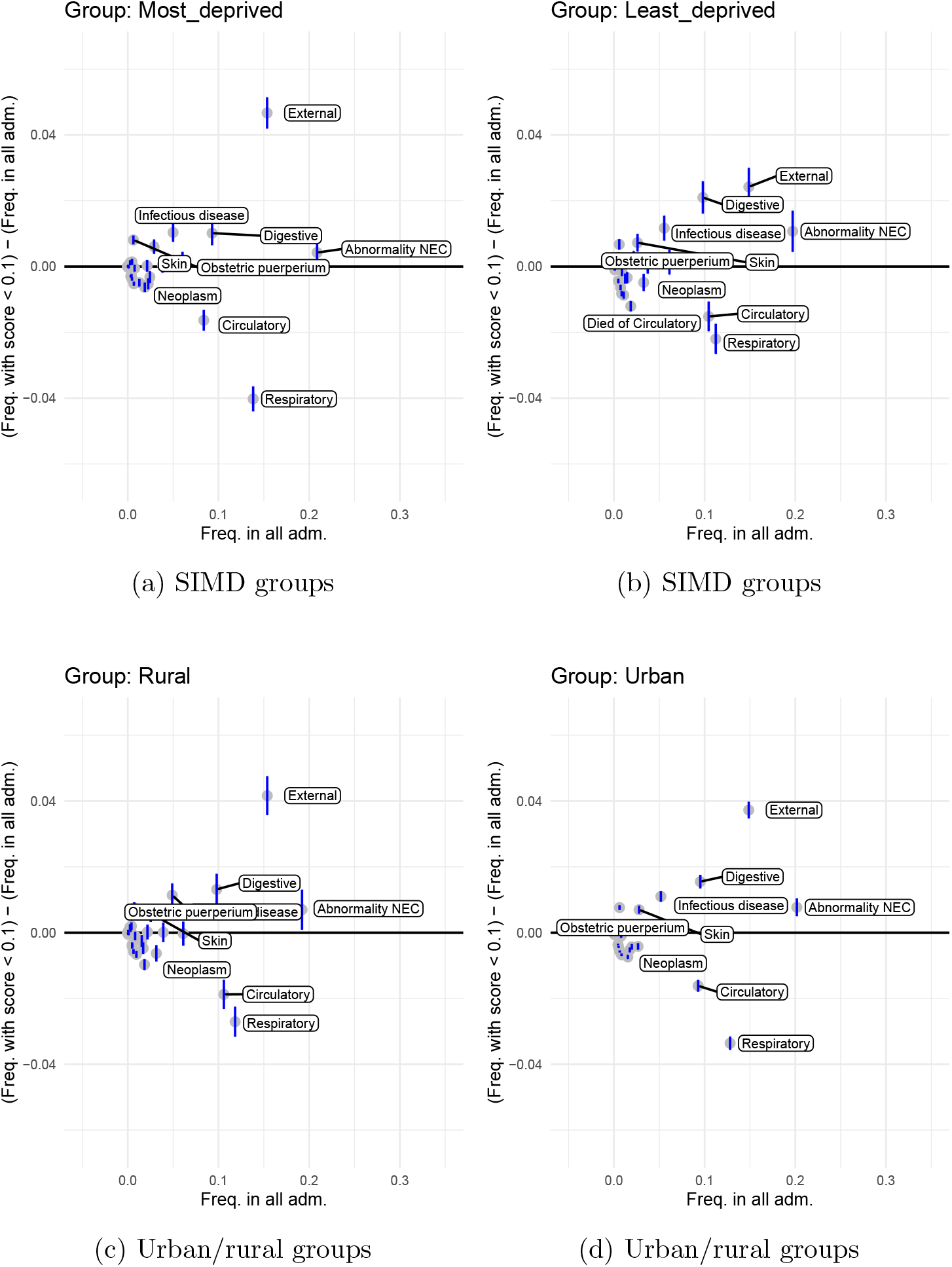
Decomposition of false negatives within SIMD and urban/rural groups. Plots consider the proportion of each admission type amongst all admissions in a group (*A*) and the proportion of each admission type amongst admissions in the group with SPARRA score *<* 10% (*B*) and plots show (*A*) against (*B*) − (*A*). Points above the line *y* = 0 correspond to admissions which are disproportionately poorly identified by SPARRA score (strictly the criterion *Ŷ <* 0.1)., Blue vertical lines show pointwise 95% confidence intervals. Upper plots show the proportion of (unpredicted) admissions due to each cause; lower plots show the proportion of deaths due to each cause. Distinctive points are labelled.

**Figure 14:**
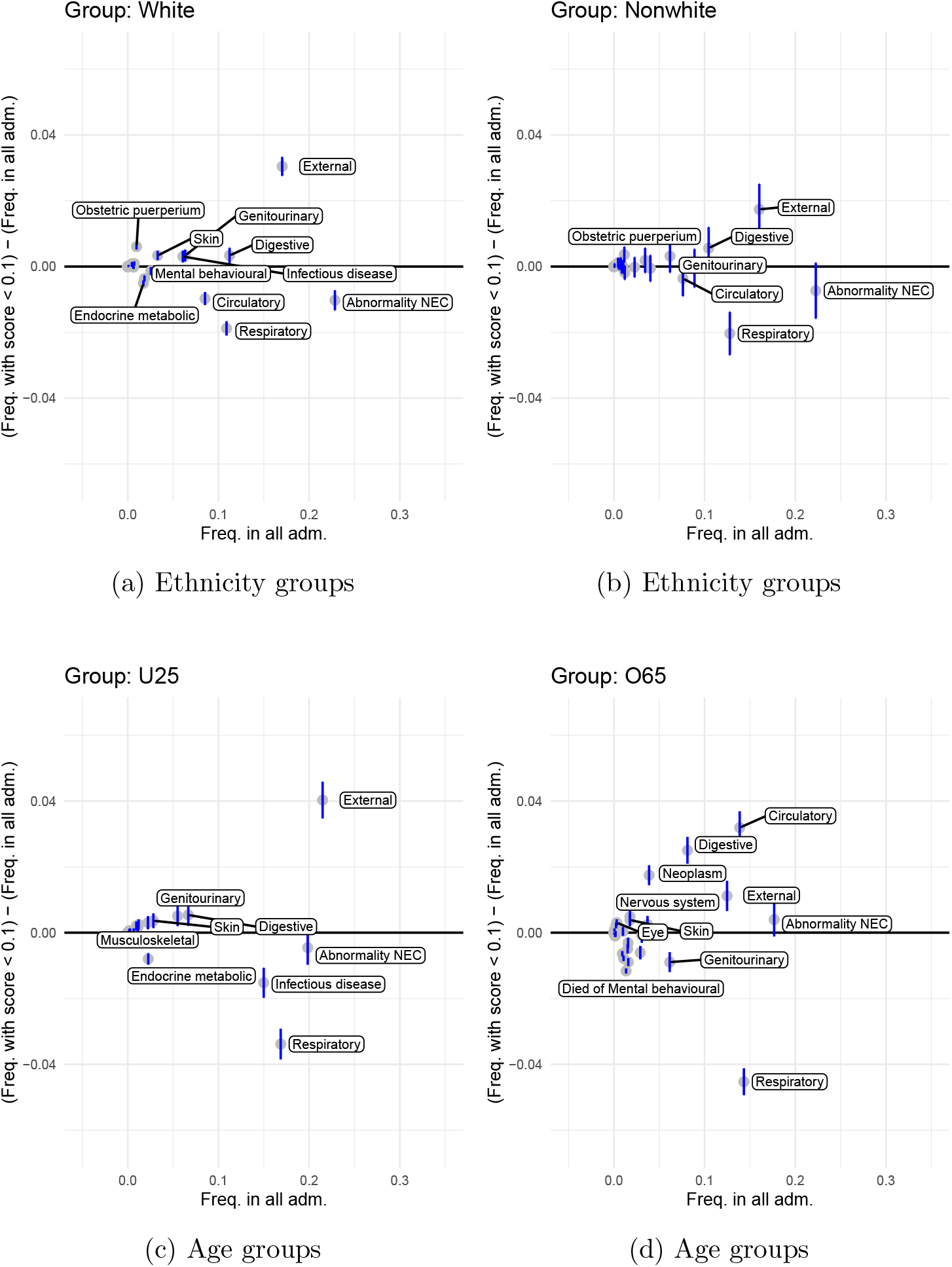
Decomposition of false negatives within ethnicity and age groups. Plots consider the proportion of each admission type amongst all admissions in a group (*A*) and the proportion of each admission type amongst admissions in the group with SPARRA score *<* 10% (*B*) and plots show (*A*) against (*B*) − (*A*). Points above the line *y* = 0 correspond to admissions which are disproportionately poorly identified by SPARRA score (strictly the criterion *Ŷ <* 0.1)., Blue vertical lines show pointwise 95% confidence intervals. Upper plots show the proportion of (unpredicted) admissions due to each cause; lower plots show the proportion of deaths due to each cause. Distinctive points are labelled.

**Figure 15:**
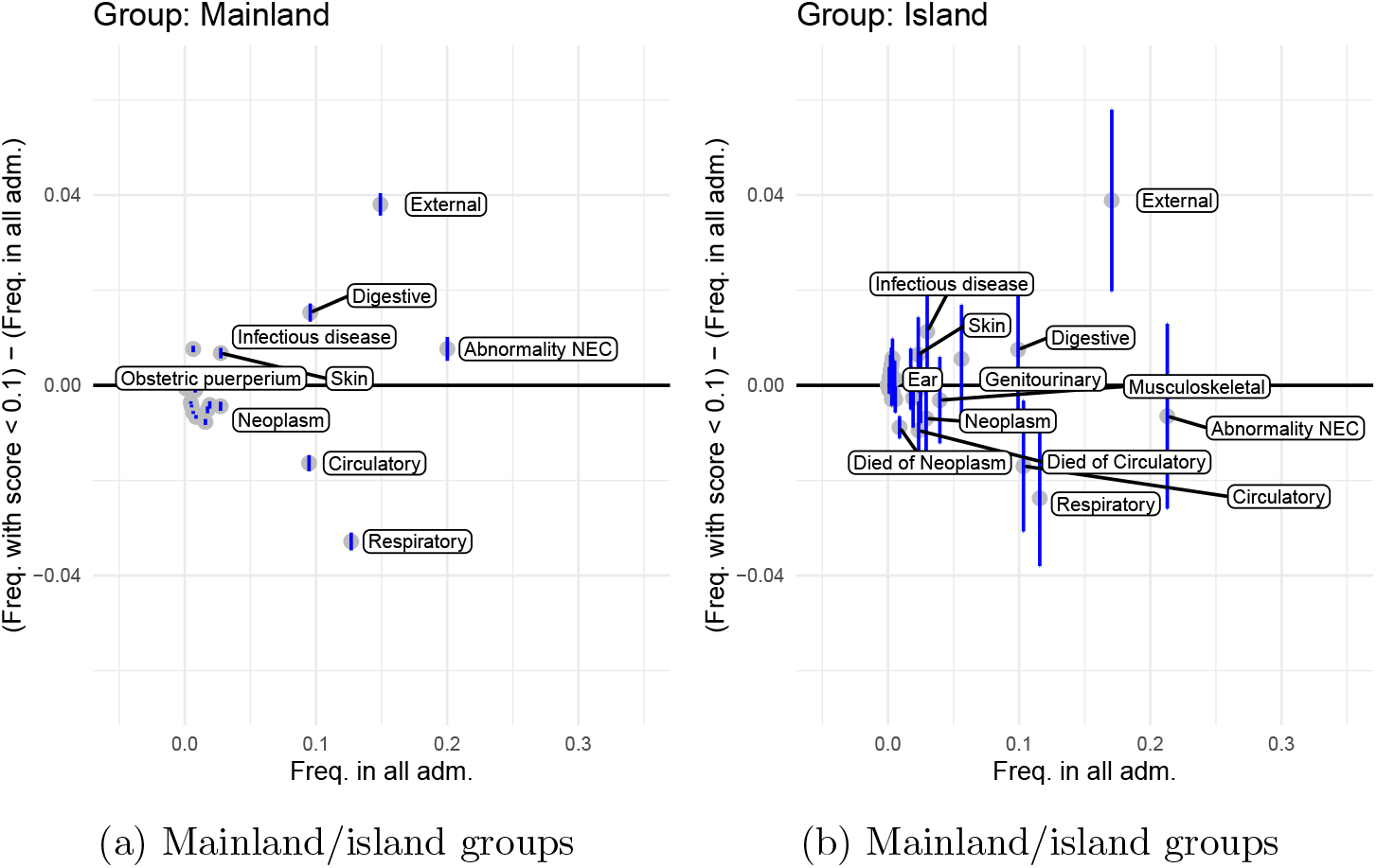
Decomposition of false negatives within mainland/island groups. Plots consider the proportion of each admission type amongst all admissions in a group (*A*) and the proportion of each admission type amongst admissions in the group with SPARRA score *<* 10% (*B*) and plots show (*A*) against (*B*) − (*A*). Points above the line *y* = 0 correspond to admissions which are disproportionately poorly identified by SPARRA score (strictly the criterion *Ŷ <* 0.1)., Blue vertical lines show pointwise 95% confidence intervals. Upper plots show the proportion of (unpredicted) admissions due to each cause; lower plots show the proportion of deaths due to each cause. Distinctive points are labelled.

## 4 Discussion

Our work indicates differential patterns of predictive performance for SPARRA across demographically defined groups in Scotland. For this purpose, we considered groups were defined by variables that are used as input for SPARRA (age, sex and deprivation), as well as other additional information that is not explicitly used when calculating the risk score (ethnicity and mainland/island or urban/rural residence status). The strength and direction of the differences varied across groups, and depending on the chosen performance metric. Moreover, our analysis also suggests that the interpretation of direct between-group comparisons is not always straightforward as the observed differences may be affected by other variables, not only on those used to define the groups. In combination, these findings highlight the importance of using a wide range of metrics, as well as adjusted comparisons (e.g. counterfactual) if the aim is to obtain a comprehensive characterisation for the performance of a risk score.

The differences revealed by our analysis do not indicate a problem with the score itself. However, we consider that it is important for practitioners and patients to be aware of these expected differential outcomes when using SPARRA. Although our performance metrics derive from the idea of algorithmic ‘fairness’ between groups, we claim that we should not generally aim to change the SPARRA score so as to eliminate between-group disparities. To do so would require a change to the objective function for SPARRA away from population-level accuracy, which necessitates a sacrifice in overall performance [30, 29, 36, 37, 38]. In other words, to eliminate disparity, we would need to trade off with the score being generally less able to predict EAs. It can even be impossible to guarantee fairness constraints (in our case, interpretable as equivalence of error rates) in some cases [36], and it is often not possible to simultaneously satisfy multiple reasonable conceptions of fairness [39]. We consider that there is little to be gained from this trade-off against overall performance, in that equivalence of error rates between groups have no obvious advantage to population well-being.

The SPARRA score is well-calibrated in essentially all groups (Supplementary Section S3.9), indicating that it is performing well at the task to which it was trained. If a practitioner aims to identify a set of individuals most likely to have emergency hospital admissions from a mixture of such groups, they should therefore identify the set of individuals with SPARRA scores exceeding some threshold, regardless of group status (Supplementary Section S3.3). Variation in error rates despite good calibration can be thought of as arising from differences in risk score distributions between groups. We argue that it is worth looking beyond calibration in this way when considering the performance of a risk score since, as discussed in the introduction, it is likely that the use of a risk score will involve decisions based on thresholds.

This study looked exclusively at routinely collected historical data, which may be subject to a variety of observational biases [40, 41]. In particular, interpretation of ethnicity data is complex due to non-random missingness (Supplementary Section S3.10. Furthermore, our work did not consider how SPARRA is integrated into the healthcare system and did not involve the analysis of healthcare decisions (e.g. primary care interventions informed by SPARRA). As such, we are not able to identify disadvantaged groups or inequity in healthcare provision, nor recommend any changes to the distribution of healthcare resources in Scotland or to the way the SPARRA score is used. However, our findings may facilitate ongoing analyses in these areas.

Our analysis is helpful for understanding and using the SPARRA score and provides insights into the epidemiology of emergency admissions in Scotland. In a general sense, we demonstrate that fairness metrics are a useful way to look at patterns of errors in a risk score.

## Data Availability

All our analysis code and data to draw figures are publically available on GitHub, where we also provide the raw code for our Shiny application and R package.

https://CRAN.R-project.org/package=SPARRAfairness

https://github.com/Public-Health-Scotland/sparra-fairness-dashboard

## Code and data availability

Our R package, SPARRAfairness implements a suite of functions used to analyse the behaviour and performance of the score, focusing particularly on differential performance over demographically defined groups. It includes utility functions to plot receiver-operator-characteristic, precision-recall and calibration curves, draw stock human figures, estimate counterfactual quantities without re-computing risk scores and simulate a semi-realistic dataset. We have provided a vignette to demonstrate how to calculate and plot a range of performance metrics for a clinical risk score across demographic groups, where we have simulated semi-realistic data for 10,000 individuals. The package includes all data necessary to reproduce the figures in this paper.

We published our complete analyses (for SPARRAv3 and SPARRAv4) in a Shiny web application [42] (available at https://github.com/Public-Health-Scotland/sparra-fairness-dashboard). This enables interactive browsing of results and we intend it to be usable by primary care practitioners and the public to enable informed decision-making.

## Ethics statement

This study and the use of patient-level EHR were approved by the Public Benefit and Privacy Panel for Health and Social Care (study number 1718-0370). Approval evidenced in application outcome minutes for 2018/19 can be viewed at https://www.informationgovernance.scot.nhs.uk/pbpphsc/application-outcomes/).

Data access was also approved by the PHS National Safe Haven through the electronic Data Research and Innovation Service (eDRIS) and the Public Benefit and Privacy Panel (PBPP) (study number 1718-0370). All studies have been conducted in accordance with information governance standards; data had no patient identifiers available to the researchers.

## Acknowledgements

We thank the Alan Turing Institute, PHS, the MRC Human Genetics Unit at the University of Edinburgh, Durham University and Health Data Research UK for their continuous support of the authors. JL, CAV, LJMA, JI, SR were supported by the Health Foundation, an independent charity committed to bringing about better health and health care for people in the UK. IT, JL, CAV, LJMA were supported by the Wave 1 of The UKRI Strategic Priorities Fund under the EPSRC Grant EP/T001569/1 and EPSRC Grant EP/W006022/1, particularly the “Health & Medical Sciences” theme within those grants & The Alan Turing Institute. IT, JL, CAV, LJMA were also supported by Health Data Research UK, an initiative funded by UKRI, the Department of Health and Social Care (England), the devolved administrations, and leading medical charities. LJMA was partially supported by a Health Programme Fellowship at The Alan Turing Institute. The funders had no role in the study design, data collection, analysis, decision to publish, or preparation of the manuscript.

## Contributions

The authors note that this project’s success was entirely contingent on close cooperation between the Alan Turing Institute and PHS. All author contributions were significant and essential to the completion of this work. Author contributions were as follows: IT, JL, CAV, LJMA, JI, SR: have made substantial contributions to the conception and design of the work, and the acquisition, analysis, and interpretation of data. IT, JL: drafted the initial work and had full access to data. CAV, LJMA, JI, SR: critically revised it for important intellectual content. KB, RP, EL, SR, JI, JL: managed data access and disclosure. KB, JI, RP, SR: provided public health input. IT, JL, CAV, LJMA, JI, SR, RP, KB: gave final approval of the version to be published.

Computing for this project was performed on the Scottish National Safe Haven (NSH), supported by the eDRIS, and the Edinburgh Parallel Computing Centre (EPCC), based at The University of Edinburgh. The authors would like to acknowledge the support of the eDRIS Team (PHS) and the NHS National Services Scotland (NSS) for their involvement in obtaining approvals, provisioning and linking data and the use of the secure analytical platform within the National Safe Haven.

For the purpose of open access, the author has applied a Creative Commons Attribution (CC BY) licence to any Author Accepted Manuscript version arising from this submission.

## Conflicts of interest

The authors declare no conflicts of interest.

**S1 Supplementary Tables**

**S2 Supplementary Figures**

## S3 Supplementary Notes

### S3.1 Notation

We firstly consider ‘covariates’ which are attributes of individuals. We differentiate covariates as follows:

*U* : all covariates; everything we know about an individual. Age, sex, SIMD, previous hospital activity data, prescriptions, ethnicity, urban-rural status. Does not include: specific postcode, marital status, smoking status, LGBT status, self-ID gender;

*X ⊂ U* : covariates used in SPARRA; age, sex, SIMD, previous hospital activity data, prescriptions. Does not include ethnicity, mainland-island status or urban-rural status;

*Z* = *U \ X* : all covariates *not* in *X*;

*A ⊂ X* : age, sex, SIMD; covariates whose effect we will attempt to either isolate ‘adjust away’.

We define the outcome value *Y* for each individual as a Bernoulli random variable indicating whether that individual was admitted to hospital in the following year (1 if admitted, 0 if not). We model covariate and outcome values (*U, Y*) for each individual as independent and identically distributed random variables with distribution (*U, Y*) *∼ 𝒟* . All probabilities and expectations are over 𝒟 unless otherwise specified. We view the SPARRA score as a fixed deterministic function *Ŷ*= *Ŷ*(*X*) *∈* (0, 100).

In general, fairness metrics concern *a decision rule made on the basis of a risk score* rather than *the risk score itself* (e.g. [43, 37, 44]). Our effective decision rule is to take some action if the event *{Ŷ ≥ c}* occurs, and not take that action otherwise. We will be concerned with individuals in various groups defined by values of *U* . We will generally use the notation *G* to indicate group membership (e.g., male sex, non-white ethnicity, island postcode). We presume *G* can be derived from *U*, so we will use *G*(*U*) where appropriate.

We will denote observations in our true data by *{*(*u*_*i*_, *ŷ*_*i*_)*}, i ∈* 1 … *n*, with *u*_*i*_ associated with *x*_*i*_, *z*_*i*_, *g*_*i*_.

### S3.2 The SPARRA score

Our study uses the third version of the SPARRA score. The score comprises four logistic regression models, each used on a subgroup of the Scottish population: FE (‘Frail elderly’ cohort; individuals aged *≥* 75), LTC (‘Long-term conditions’; individuals aged 16-75 with prior healthcare system contact), YED (‘Young emergency department’; individuals aged 16-55 who have had at least one A&E attendance in the previous year) and U16 (‘under-16’; individuals aged *<* 16). These groups cover approximately 80% of the Scottish population. For individuals present in both the LTC and YED subgroups, the maximum of the two risk prediction scores is reported. Input features include age, sex, an index of socioeconomic deprivation (using the deciles of the 2016 Scottish Index of Multiple Deprivation (SIMD) [45] as a geographic-based proxy), as well as information about long-term conditions, past hospital activity data and prescriptions. All features were derived from national electronic healthcare records (EHR) databases held by PHS, considering information up to three years prior to the prediction time cutoff (except for long-term conditions, which are extracted from historic records since 1981). SPARRAv3 scores were calculated by PHS and provided as input for our analysis. SPARRAv3 predicts emergency in-patient admission within 12 months from a given time cutoff. When training the model, individuals who died before the time cutoff or within 12 months after were excluded. In this analysis, rather than excluding individuals who died after the time cutoff, we considered a composite outcome of death or EA in the 12 months following the time cutoff. Since death indicates a poor health outcome that would otherwise be excluded from analysis, we can broadly consider SPARRAv3 as a tool for predicting abrupt breakdowns in health more generally [15].

We also repeated the analysis using SPARRAv4 [15], which is expected to be deployed in Scotland in 2024. SPARRAv4 was trained using more recent versions (2013-2018) of the same input data sources as SPARRAv3 and using more complex machine learning methods (e.g. random forests, gradient-boosted trees). This improved accuracy over SPARRAv3, though scores remained broadly similar [15]. To avoid duplication, this manuscript only presents results based on SPARRAv3. Other results can be explored on our interactive web application.

### S3.3 Choice of patients on whom to intervene

Suppose we are a practitioner with a set of patients for whom we have a well-calibrated risk score giving their probability of emergency hospital admission in the coming year. We have the resources to intervene on only a fixed number of patients, and we wish to choose these patients so as to find as many as possible who would go on to have an admission.

Intuitively, we would expect that the best way to choose this subset of patients would be to target those patients with risk scores exceeding some threshold. We will show that this intuition is correct, and that, although the risk score may exhibit different behaviour across subgroups of patients, we are best not to change this action threshold across groups.

When we say our risk score *Ŷ*= *Ŷ*(*X*) is well-calibrated, we mean that it satisfies:

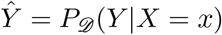

that is, which accurately estimates the risk of admission for an individual with covariates *X* = *x*.

Denote by 𝔛 the domain of *X*, corresponding to the range of values covariates can take, where *X* is a random variable with measure *μ* (so for a set of potential covariate values *X*^*′*^, *μ*(*X*^*′*^) is the frequency at which patients have covariates in *X*^*′*^). We wish to choose a range of possible covariate values on which we will intervene. Due to cost constraints, we can only intervene on a fixed proportion of individuals.

Thus we wish to choose a region Γ *⊂ 𝔛* with fixed measure *γ* = *μ*(Γ), optimising some objective. If we simply wish to choose Γ so as to maximise the objective *P* (*Y* |*X ∈* Γ), then we should choose Γ = *{x* : *Ŷ* (*x*) *> c}* for some *c*, since if Γ^*′*^*≠* Γ but *μ*(Γ) = *μ*(Γ^*′*^) and *μ*(Γ^*′*^ *\* Γ) *>* 0 we have

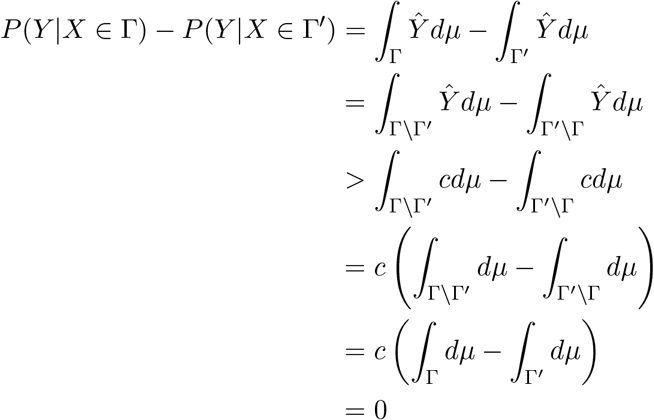

that is, if a practitioner wishes only to choose a subset of patients on whom to intervene such that they have the highest expected frequency of admissions, they should simply choose those patients for whom the SPARRA score exceeds some threshold. Intuitively, if we start with a group of individuals for which the SPARRA score exceeds a threshold *c*, and we change the group by removing someone and replacing them with someone else, then since the person we removed has a SPARRA score greater than *c* and the person who replaced them has a score less than *c*, we are necessarily reducing the expected number of people we find who will eventually be admitted.

Importantly, this is independent of any group structure: if we have the option of varying the threshold between groups, if we simply aim to ‘catch’ as many individuals as possible with *Y* = 1, we should use an identical threshold on *Ŷ* in all groups. We consider groupings defined by values of a random variable *G*. Because the score is well-calibrated for all groups, we have (essentially) *P* (*Y* |*G* = *g, Ŷ* (*X*) = *y*) = *P* (*Ŷ*(*X*) = *y*) = *y*, so *Ŷ****⊥*** *G*|*X*; or equivalently, *G* does not contribute further information to knowledge of the probability of *Y* = 1 once we know *X*. Hence the optimality of Γ does not depend on our groupings, and a practitioner aiming to target patients so as to target as many admissions as possible should target individuals with score in excess of a given threshold, where the threshold is identical over groups.

If we consider use of the same thresholds *c* in two groups *G* = *g* and *G* = *g*^*′*^ does not guarantee that *P* (*Y* = 1|*G* = *g, Ŷ< c*) = *P* (*Y* = 1|*G* = *g*^*′*^, *Ŷ < c*); that is, perfect calibration does not guarantee that FOR is identical between groups.

### S3.4 Score distribution

#### S3.4.1 Demographic parity

We firstly assess demographic parity [46, 3 8]: the distribution of scores across groups, termed *DP*, defined as

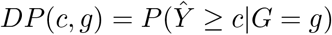

across cutoffs *c* and groups *g* . We use the standard (uniformly consistent) CDF estimator:

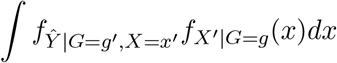

#### S3.4.2 Counterfactual fairness

We then compute counterfactual fairness between groups, which can be thought of as describing differences in distribution of score due only to the effect of group on (some of) age, sex, and deprivation and directly on the score. We do this by considering a hypothetical individual who resembles a typical member of a given group, except that they are a member of another different group. This requires the specification of a system of causality relating covariates, group, and predicted score. We use the following causal model:

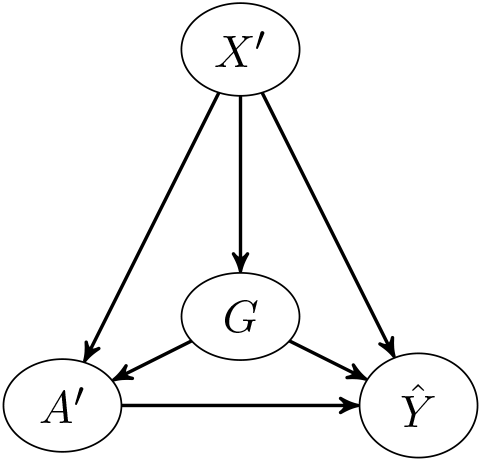

In this graph, vertex *G* denotes group (e.g. urban/rural, older/younger). Vertex *A*^*′*^ denotes (*A \ G*); that is, age, sex and SIMD, except any of these which determine *G*. Vertex *X*^*′*^ denotes (*X \* (*A ∪ G*)); that is, covariates in SPARRA score excepting age, sex, SIMD and variable determining *G*. Vertex *Ŷ* denotes the prediction from SPARRA score. The distribution of *background variable X*^*′*^ is its marginal distribution in 𝒟.

The *structural equation* for vertex *V* with parents *P* is denoted 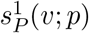 and is given by 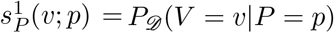, substituting densities for probabilities where appropriate. For instance, the structural equation for *A* is 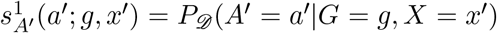 . The superscript ‘1’ identifies the structural equation with this causal graph as opposed to later causal graphs. We compare the counterfactual values, which we term *DPC*:

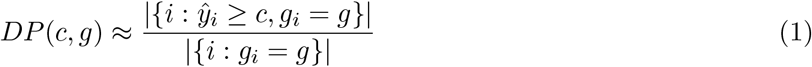

between cutoffs *c*, and for either *g* ^*′*^ = *g* or *g* ^*′*^*≠ g* . We compute values (2) in three steps [30, 31]:

1. Compute the posterior distribution 𝔛 ^*′*^ *∼* (*X*^*′*^|*G* = *g*); that is, the distribution of everything other than age, sex, SIMD for members of *g*.
2. Delete the edge from *X*^*′*^ to *G* and set *G* = *g*^*′*^; that is ignore the influence of non-age, sex, SIMD covariates on group status, and presume we are dealing with a member of *g*^*′*^,
3. Compute the joint distribution (𝔛, *A* ^*′*^) with density at *x*^*′*^, *a*^*′*^ given by

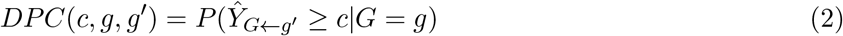

that is, compute the distribution of non-group covariates for a person whose group is *g*, but for which age, sex and SIMD are distributed as though they were a random member of the population with *Ŷ < c*,

1. Set random variable (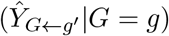) with density at *y* given by

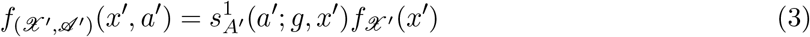

that is, compute the distribution of predicted score of a person for which all non-age, sex, SIMD covariates are distributed as though they were in group *g*, but they are in group *g*^*′*^ and have corresponding values of age, sex, and SIMD.

We sample from the distribution of 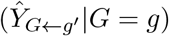 as follows:

1. Choose a random individual with *G* = *g*, and note their values *x*^*′*^ of *X*^*′*^
2. Choose a random individual with *G* = *g*^*′*^ for whom *X* = *x*^*′*^
3. Record the value of *Ŷ* for this individual as a sample from 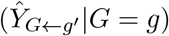

To see why this works, we note that we are sampling a random individual with *X* = *x*^*′*^ and *G* = *g*^*′*^; hence the distribution of *Ŷ* we obtain in step 3 conditional on our choice of *x*^*′*^ in step 2 is identical to that of (*Ŷ* |*G* = *g*^*′*^, *X* = *x*^*′*^) and unconditional on *x*^*′*^ is

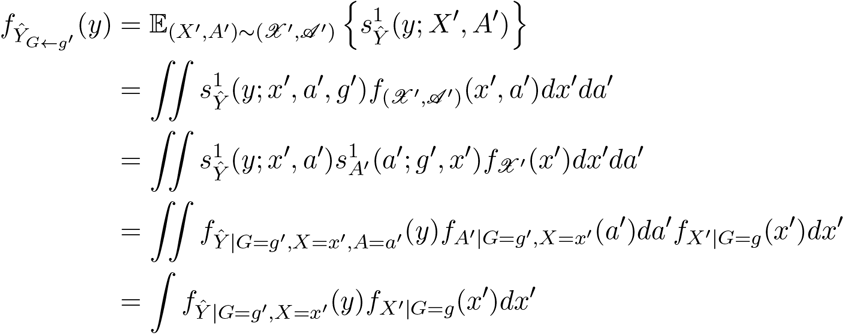

Working from the definition of 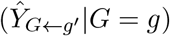 we have

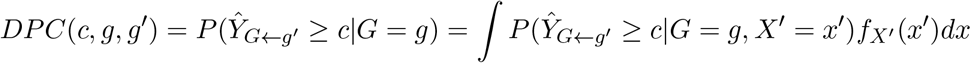

as required. For group *g*, rather than choosing a random individual in step 1, we run through all samples *i* with *g*_*i*_ = *g*, producing |*{i* : *g*_*i*_ = *g}*| corresponding samples of 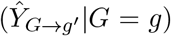 . We estimate quantity 2 analogously to the estimate (1) with the estimates of 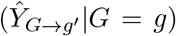 in place of the *y*_*i*_. We provide a general implementation of this procedure in our R package.

Formal counterfactual fairness [30] requires the identity:

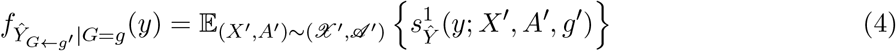

to hold for all values *x*^*′*^ and *g*; we only compare the mean quantities:

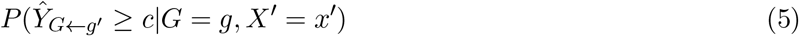

where 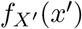 is the marginal density of *X*^*′*^ in the common distribution of (*U, Y*).

The comparison of counterfactuals between groups amounts to assessment of the difference in distribution of *Ŷ* attributable only to effects of group on *A*^*′*^. Broadly, we are interested in the two classes of causes of variation in *Ŷ* : *A*^*′*^ and *X*^*′*^. We view *X*^*′*^ (hospital activity data, prescriptions, long term conditions) as ‘underlying’ causes of variation in *Ŷ*, where variables in *A*^*′*^ act as ‘modulators’ to the score. We model this with an edge *X*^*′*^ to *A*^*′*^.

We note that groups *G* are associated with different posteriors over *X*, so we model an edge *X*^*′*^ to *G*. We wish to include the effects of group *G* on the ‘modulation’ of the effect of *X*^*′*^ on *Y* through *A*^*′*^ so include an edge from *G* to *A*^*′*^, but discount the variation in posteriors in ‘legitimate’ covariates *X*^*′*^. The counterfactual 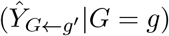 achieves this by assessing the difference in *Ŷ* between that arising from the full causal graph and the causal graph with the *X*^*′*^ *→ G* edge removed, while choosing the background variable *X*^*′*^ to be distributed as though *G* were *g* in both cases.

### S3.5 False negatives

#### S3.5.1 Raw False Omission Rate

We characterise ‘false negative’ errors using false omission rate (FOR), termed *FOR* [27, 44]. We compare the values of

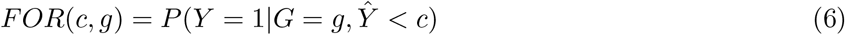

across cutoffs *c* and groups *g*. The events *Y* = 1, *Ŷ < c* indicate in a sense that the prediction *Ŷ < c* was incorrect. The FOR indicates a measure of how often individuals in a group predicted *not* to be admitted were in fact admitted. We estimate FOR consistently as

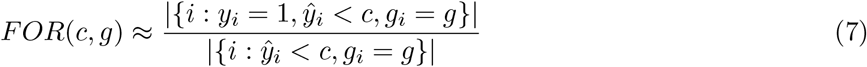

### S3.5.2 Adjusted FOR

We also consider FOR ‘adjusted’ for variables *A*^*′*^ = *{A \ G}* (age, sex, and deprivation, excluding any variables that determine *G*). We denote

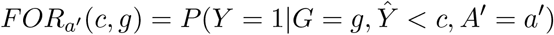

from which we may write quantity (6) as:

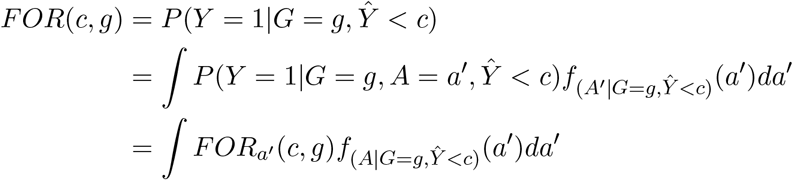

To adjust for *A*^*′*^, we replace the density 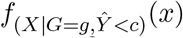 with 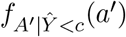, which does not depend on *g*. This generates adjusted FOR, which we term *FOR*_*A*_:

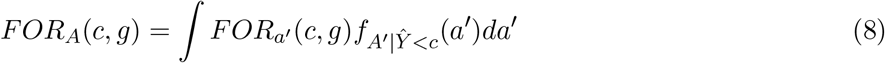

Our data discretises *A*^*′*^ (rounding age to the nearest year). Writing 𝔸 as the set of possible discrete *A*^*′*^ values, we estimate *FOR*_*A*_(*c, g*) as:

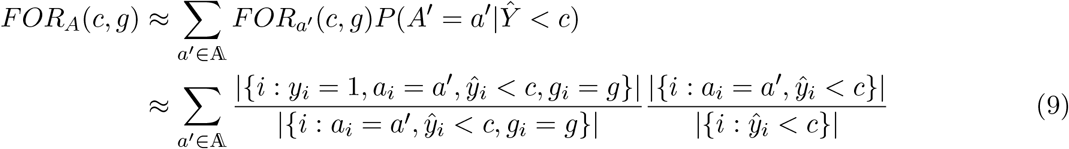

where the first term in the sum is an estimate of *FOR*_*a*_^*′*^ (*c, g*) and the second is an estimate of *P* (*A*^*′*^ = *a*^*′*^|*Ŷ < c*). The second approximation is consistent.

### S3.5.3 Adjusted FOR as counterfactual

The quantity *FOR*_*A*_(*c, g*) can be viewed as a comparison of counterfactual quantities

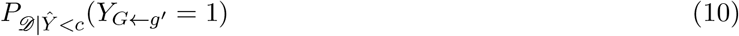

across groups *g*, under the causal specification as follows:

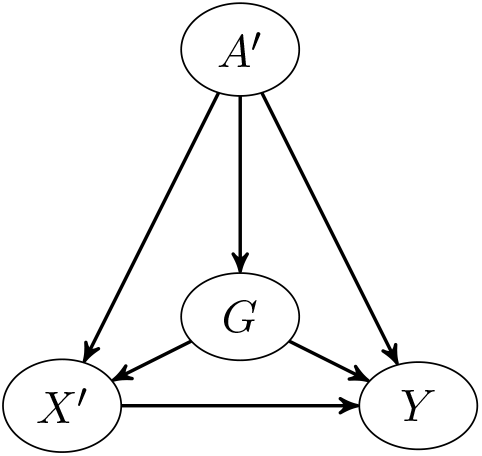

Note *X*^*′*^ and *A*^*′*^ are swapped from their positions in the previous causal graph. Vertices *G, X*^*′*^ and *A*^*′*^ have the same meanings as in Section S3.4.2; vertex *Y* indicates outcome; that is, admission or non-admission in coming year. The *background variable A*^*′*^ has distribution given by the marginal of *A*^*′*^ in 𝒟|*Ŷ < c*. Similarly to the previous causal graph, we denote the *structural equation* of vertex *V* with parents *P* as 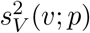 given by 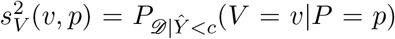 . In this case, however, probabilities in the background variables and structural equations are with respect to 𝒟|*Ŷ < c* rather than 𝒟 .

To see the equivalence of quantities (8) and (10), note that the analogous procedure to counterfactual fairness above computes 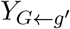 as:

1. Denote by *A* ^*′*^ the distribution of *A*^*′*^|*Ŷ < c*, or equivalently the marginal of *A*^*′*^ in *D*|*Ŷ < c*; that is, the distribution of age, sex and SIMD over the whole population.
2. Delete the edge from *A*^*′*^ to *G* in the graph and set *G* = *g*,; that is ignore the influence of non-age, sex, SIMD covariates on group status, and presume we are dealing with a member of *g* whose values of *A*^*′*^ are sampled as though they were a random member of the population with *Ŷ < c*.
3. Compute the joint distribution (𝔛, *A* ^*′*^) with density at *x*^*′*^, *a*^*′*^ given by 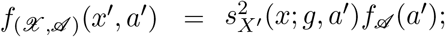 that is, compute the distribution of non-group covariates for a person whose group is *g*, but for which age, sex and SIMD are distributed as though they were a random member of the population with *Ŷ < c*,
4. Now 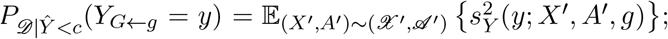 that is, we compute the distribution of predicted score of a person for which age, sex, and SIMD are distributed as though they were a random member of the population, but for which their group is *g*.

Now

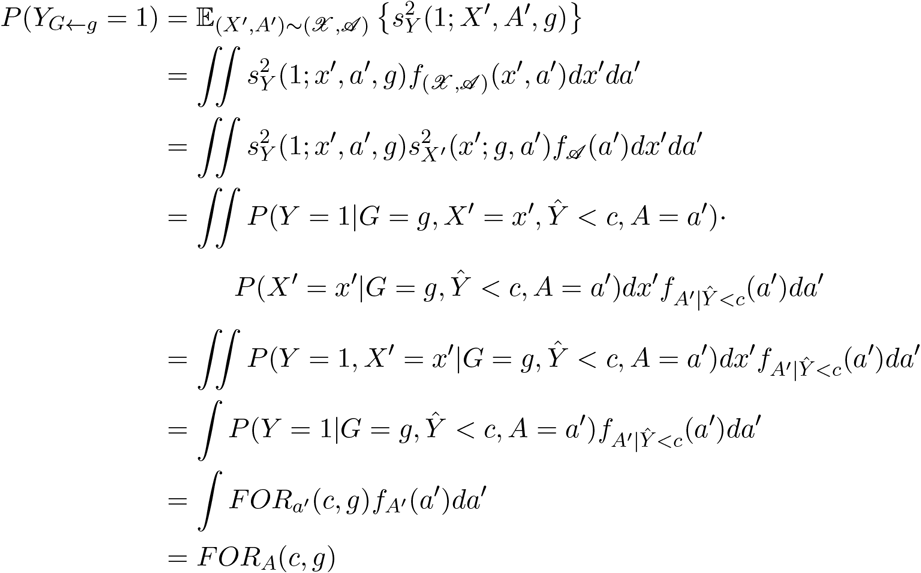

In contrast to *DPC*, the counterfactual in this case adjusts away for the effect of group *G* on demographics *A*^*′*^ while including the effects of *G* on *Y* . Correspondingly, the causal graph in this case has reversed positions of *X*^*′*^, *A*^*′*^ compared with the causal graph in Section S3.4.2

An alternative measure of false negatives would be to measure the false negative rates directly; for instance, using *FNP* which compares

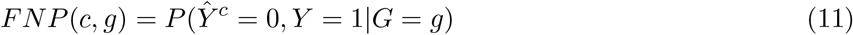

between groups *g* and across cutoffs *c*. We chose *FOR* ahead of *FNP* because the population of individuals of interest to GPs is better represented by the individuals for which *G* = *g, Ŷ* ^*c*^ = 0 (the conditional population in *FOR*) than the population with just *G* = *g* (the conditional population in *FNP*), in that the former represents individuals with *G* = *g* judged as low-risk by SPARRA. We compare *FNP* (*c, g*) between groups *g* in the Supplementary Figures and in the Shiny app associated with this manuscript.

### S3.5.4 Outcome disparity

Individuals in the set of the numerator of the *FOR*(*c, g*) estimate (equation 7) were all admitted to hospital (*y*_*i*_ = 1) despite being ‘predicted not to’ (ŷ_*i*_ *< c*). Most individuals admitted to hospital have a recorded primary diagnosis, coded according to the ICD10 criteria [47]. We considered the distribution of the first letter of these ICD10 codes across individuals, giving a breakdown of admission causes amongst admitted individuals with ŷ_*i*_ *< c* (Supplementary Table 3).

Denote admission causes by a random variable *W* (with observations *w*_*i*_). For an admission cause *w*, group *g*, and fixed score cutoff *c* = 0.1, we make the estimates

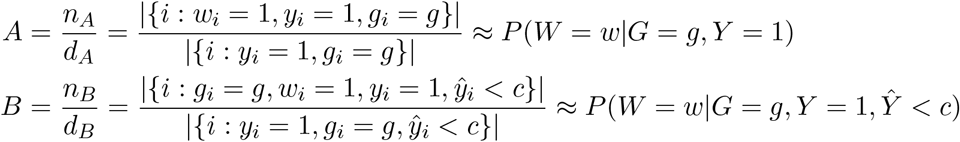

and plot *A* against *B* − *A* (Figures 3 and similar). We estimate standard errors of *B* − *A* as:

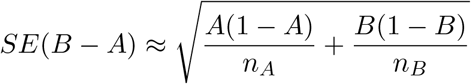

and confidence intervals through these standard errors.

### S3.6 False positives

We characterise ‘false positive’ errors analogously to false-negative errors using false-discovery rate parity, termed FDRP [27], defined as

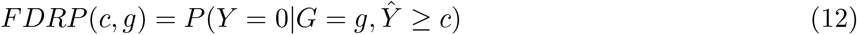

and again compute the ‘adjusted’ version of this quantity, defined as:

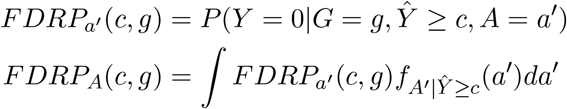

which can be viewed as a comparison of counterfactual quantities

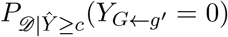

across groups *g*, under the causal specification in Section S3.5, noting that probabilities are with respect to the distribution (𝒟|*Ŷ ≥ c*) rather than (𝒟|*Ŷ < c*). We estimated these in an analogous way to *FOR*(*c, g*) and *FOR*_*A*_(*c, g*) (equations (7) and (9)

### S3.7 Overall accuracy

#### S3.7.1 ROC curves

We firstly analyse group-level accuracy in each group using discrimination (area under receiver-operator characteristic, AUROC) and calibration. The AUROC (also called the C-statistic) estimates

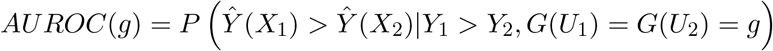

over groups *g*, where (*U*_1_, *Y*_1_) = (*X*_1_, *Z*_1_, *Y*_1_) and (*U*_2_, *Y*_2_) = (*X*_2_, *Z*_2_, *Y*_2_) have the common distribution of (*U, Y*). That is, the AUROC estimates the probability that the score for a randomly chosen admitted individual exceeds the score for a randomly chosen non-admitted individual. We use the (consistent) estimator:

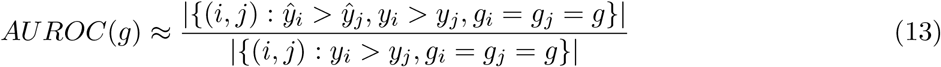

### S3.7.2 Calibration curves

We secondly analyse group-level accuracy by means of calibration curves (also called reliability diagrams) [34] in which for a range of cutoffs *c* we compare the difference of *P* (*Y* |*Ŷ* = *c*) from *c*: that is,whether individuals with a score of *c* are admitted in on average *c*% of cases.

We consider the intervals 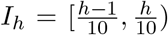 for *h ∈* 1 : 10. For each interval we make the (consistent) estimates:

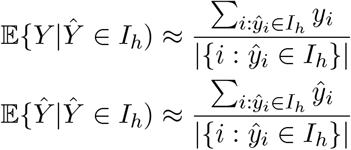

with confidence intervals and standard errors estimated using the usual asymptotic confidence interval of a proportion. We plot these estimates directly for comparison.

### S3.8 Other metrics of fairness

We also compute the following metrics for group fairness, available on our Shiny app. These are all compared between groups *g* at a range of cutoffs *c*.

*FPP* (*c, g*) : False positive parity: *P* (*Ŷ* = 1, *Y* = 0|*G* = *g*)

*FPRP* (*c, g*) : False positive rate parity: *P* (*Ŷ* = 1, *Y* = 0|*G* = *g*)

*RP* (*c, g*) : Recall parity: *P* (*Ŷ* = 1|*G* = *g, Y* = 1)

*IRP* (*c, g*) : Inverse recall parity: *P* (*Ŷ* = 0|*G* = *g, Y* = 0)

*FNP* (*c, g*) : False negative parity: *P* (*Ŷ* = 0, *Y* = 1|*G* = *g*)

*FNRP* (*c, g*) : False negative rate parity: *P* (*Ŷ* = 0|*G* = *g, Y* = 1)

All are estimated using consistent estimators analogous way to that used for *FOR*(*c, g*) (equation (7)). We provide general code to estimate all metrics in our R package.

### S3.9 Discrimination and calibration

In comparison with the overall cohort, the largest differences in discrimination (as measured by AUROC) were observed for the subgroups defined by age and ethnicity. Discrimination was stronger in individuals over 65 than in those younger than 25 (Supplementary Figure 6a), indicating that EAs are more readily predictable within this group. Although both nonwhite and white subgroups had poorer discrimination than the overall cohort (Supplementary Figure 6d), we did not observe large differences in AUROC between these subgroups. Less prominent differences were observed in all other comparisons, where AUROC within each subgroup was similar to the overall AUROC: discrimination was slightly stronger in the rural than in urban residents (Supplementary Figure 6e) and in females than in males (Supplementary Figure 6b), but differences were almost negligible between mainland and island residents (Supplementary Figure 6f).

Despite small deviations between the predicted and observed number of events, SPARRA is generally well-calibrated in the overall cohort, and within most of the subgroups considered in our analysis (Supplementary Figure 7). The largest departures from perfect calibration were observed for the subgroups defined by age and ethnicity. The estimated calibration curve suggests that SPARRA may overestimate risk for the younger group, although with a high degree of uncertainty attached (Supplementary Figure 7a). Moreover, SPARRA also appears to underestimate EA risk for both white and nonwhite subgroups (Supplementary Figure 7d).

### S3.10 Determination of ethnicity

Ethnicity data was determined for individuals in our dataset by Public Health Scotland by cross-reference of information across a range of sources. In particular, the following datasets were considered:

1. Records from the COVID-19 vaccination program from November 2021 onwards
2. Outpatient and inpatient or day case hospital records from March 2010 [48, 49]
3. Rapid Preliminary hospital Inpatient Data records from February 2020 [50]
4. Data from the COVID Case Management System from June 2020
5. Electronic Communication of Surveillance in Scotland from February 2020 [51]
6. Urgent Care Datamart from January 2011 Methodology [52, 53]

For each source of ethnicity, the CHI number (a national individual-level identifier), ethnicity identifier and date of capture were imported. The ethnicity identifier varies between data sources, so each lookup has its own mapping.

Ethnicity classifications are based on the Scottish 2011 Census categories [54] which are used as a standard across NHS Scotland.

Once ethnicity identifiers had been mapped records were removed where

1. CHI number is missing or appears malformed.
2. Ethnicity identifier could not be mapped.

All sources were joined and sorted by date of capture. Ethnicity records are considered valid for inclusion if any of the following were true:

1. The ethnicity did not come from the vaccination program
2. The ethnicity came from the vaccination program, but was added before 5th September 2022

When two or more ethnicity records were present for a given patient, the most recent ethnicity recorded for a patient was retained.

Our analysis revealed some differences between individuals with white and nonwhite recorded ethnicity. Among others, calibration was poorer for both groups than for the overall cohort, but better for white than nonwhite ethnic groups. However, the interpretation of these results is not straightforward due to high levels of non-random missingness for ethnicity.

## Notes

### Competing Interest Statement

The authors have declared no competing interest.

### Author Declarations

This study and the use of patient-level EHR were approved by the Public Benefit and Privacy Panel for Health and Social Care (study number 1718-0370). Approval evidenced in application outcome minutes for 2018/19 can be viewed at https://www.informationgovernance.scot.nhs.uk/pbpphsc/application-outcomes/). Data access was also approved by the PHS National Safe Haven through the electronic Data Research and Innovation Service (eDRIS) and the Public Benefit and Privacy Panel (PBPP) (study number 1718-0370).

